# A clinic-updated digital twin for Parkinson’s disease progression: Governed Bayesian forecasting with uncertainty-gated reporting

**DOI:** 10.64898/2026.03.19.26348807

**Authors:** Ahmed Abdelmonem Hemedan, Vladimir Despotovic, Taras O. Lukashiv, Kinza Rian, Sonja R. Jónsdóttir, Claire Pauly, Lukas Pavelka, Sibylle Béchet, Esther Ademola, Morgan Leonard, Luiz Carlos Maia Ladeira, Petr V. Nazarov, Liesbet Geris, Venkata P. Satagopam, Rejko Krüger

**Affiliations:** Transversal Translational Medicine, Luxembourg Institute of Health (LIH), 1445 Strassen, Luxembourg; Bioinformatics and AI, Department of Medical Informatics, LIH, 1445 Strassen, Luxembourg; Bioinformatics Area, Fundación Progreso y Salud (FPS), Sevilla, Spain; Parkinson Research Clinic, Centre Hospitalier de Luxembourg (CHL), 1210 Luxembourg, Luxembourg; Bioinformatics Core, Luxembourg Centre for Systems Biomedicine (LCSB), University of Luxembourg, 4362 Esch-sur-Alzette, Luxembourg; Translational Neuroscience, LCSB, University of Luxembourg, 4362 Esch-sur-Alzette, Luxembourg; Parkinson Research Clinic and Department of Neurology, CHL, 1210 Luxembourg, Luxembourg; GIGA Molecular Biology of Diseases, Université de Liège, Li`ege, Belgium; Multiomics Data Science, Department of Cancer Research, LIH, 1445 Strassen, Luxembourg; Biomechanics Research Unit, University of Liège, Liège, Belgium; Department of Mechanical Engineering, KU Leuven, Leuven, Belgium; Translational Informatics Group, LCSB, University of Luxembourg, 4362 Esch-sur-Alzette, Luxembourg

## Abstract

**Background:** Clinical digital twins hold considerable promise for forecasting disease progression, yet the question of when a model’s outputs should be withheld remains largely unaddressed. A predictive model qualifies as a governed reporting system only when it specifies the operational boundaries under which its outputs are reliable and enforces criteria for suppressing results that fall outside those bounds.

**Methods:** We present a governed Bayesian digital twin for multi-domain Parkinson’s disease (PD) progression, tracking motor function (MDS-UPDRS Part III), cognition (Montreal Cognitive Assessment, MoCA), and autonomic function (SCOPA-AUT). A monotone latent state-space model captures disease progression under four architectural constraints: non-decreasing latent severity, visit-triggered updating, full posterior uncertainty propagation, and non-causal scope. A six-rule confidence gate evaluates each forecast before release; when evidence is insufficient, the gate suppresses the output and returns a structured reason code. We evaluated the framework on the Parkinson’s Progression Markers Initiative (PPMI), a multicentre longitudinal observational study (*N*=4,628 participants; 28,185 visits), using five-fold cross-validation with independent model refits, equity analysis, and coupling-topology sensitivity assessment. The framework is available at https://gitlab.com/ahmed.hemedan/symphony-dt, with a research prototype at https://symphony-dt.com/.

**Results:** Predictive interval coverage at the 95% level ranged from 94% to 96% across all three endpoints, compared with 64–69% for linear mixed-effects baselines. The confidence gate released governed forecasts at 32.7% of visits under strict three-domain requirements, increasing to 48.1% under a validated partial-observation extension. Suppression was predominantly driven by incomplete clinical assessment (51.5%) rather than model uncertainty (0.2%), and operated equitably across sexes (Cramér’s *V*=0.049). Five of six cross-domain coupling parameters were identified from the data (sign probability ≥ 0.99; contraction ratios 0.19–0.35), with all cross-domain forecast correlations matching the directions predicted by the coupling topology. The framework’s own diagnostics localised two observation-model limitations, Prodromal motor heteroscedasticity and medication-burden sensitivity, to a single model layer and specified their resolution.

**Conclusions:** Governed silence, defined as the rule-based suppression of predictions when reliability conditions are not met, can be embedded in clinical prediction architecture, quantified as a pipeline output, and audited for equity. This work demonstrates the technical executability of governed digital twin architecture at cohort scale and provides a foundation for prospective deployment under routine clinical conditions.

## 1 Introduction

Digital twins have become a central organising concept in precision medicine [10, 11]. The premise is a patient-specific model, updated with new data, generating forecasts that support decisions. Scoping reviews document growing methodological diversity [11, 28]. Yet deployment remains rare, and the gap is structural.

A predictive model becomes a governed reporting system only when it specifies when to withhold its outputs [23]. Calibrated uncertainty quantifies interval width but does not decide whether the interval should be shown [7]. External validation extends scope but does not define operational boundaries [8, 9]. Deployment requires a reporting framework: a set of rules stating when silence— the deliberate suppression of a prediction because reliability conditions are not met—is the correct output. That framework must be auditable. It must also demonstrate that suppression does not accumulate in vulnerable subgroups.

Parkinson’s disease (PD) progression monitoring stress-tests every component of such a framework. PD drives multi-domain decline across motor, cognitive, and autonomic systems [1, 27], typically assessed via the Movement Disorder Society Unified Parkinson’s Disease Rating Scale Part III (MDS-UPDRS III) for motor function, the Montreal Cognitive Assessment (MoCA) for cognition, and the Scales for Outcomes in Parkinson’s Disease—Autonomic (SCOPA-AUT) for autonomic dysfunction. Rates differ across patients and accelerate unevenly [2]. Clinic visits are intermittent, with gaps spanning months [3]. Recorded scores shift with medication state, rater context, and timing [4]. Multi-domain assessment is incomplete at the majority of visits. A model that reports reliably under these conditions provides evidence for the broader class.

Existing PD progression models address complementary aspects of this challenge. Subtyping approaches identify trajectory clusters [2, 5]. Machine learning pipelines predict worsening events [6]. Mechanistic models encode pathway couplings [38]. Bayesian trajectory models estimate individual courses with uncertainty [18]. Each has advanced the field, yet all are evaluated by average accuracy on held-out data. For deployment, the critical question is visit-specific: can *this* forecast, for *this* patient, be reported? Answering that question requires operational reporting boundaries within the evaluation itself.

The governed twin addresses this through four architectural constraints and a six-rule confidence gate. Latent progression is non-decreasing, reflecting irreversible neurodegeneration [12]. State estimation occurs only at clinic visit times. Uncertainty propagation is integral to every output. The model scope is associational, tracking progression dynamics conditional on observed history. The six-rule gate evaluates each forecast before release. When any rule fires, the system returns a scope statement naming the reason. Figure 1 summarises the architecture.

**Figure 1:**
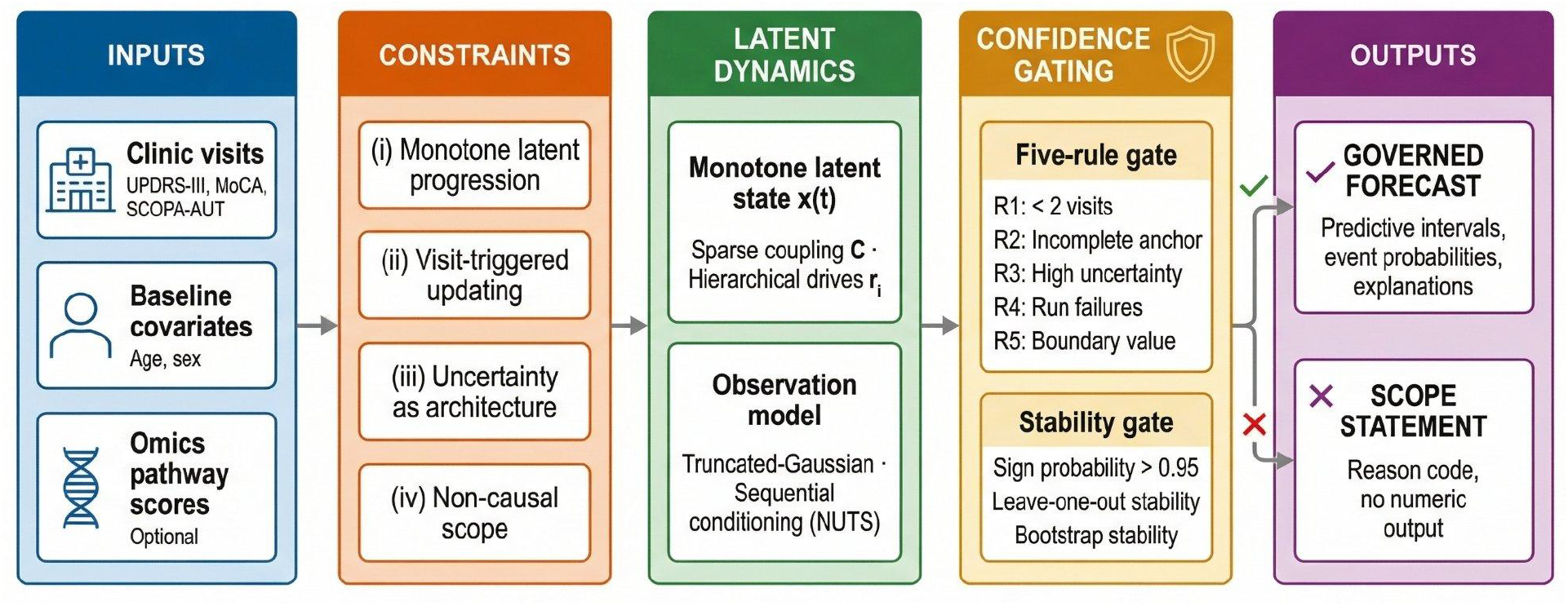
Architecture of the governed digital twin. Inputs flow through four architectural constraints into the latent dynamics and observation model. A confidence gate and a stability gate determine whether each forecast is released (governed output) or suppressed (scope statement with reason code).

Throughout this paper, a *governed* forecast has passed all gating rules and is released for clinical review. A *suppressed* forecast has failed at least one rule and is replaced by a scope statement identifying the reason.

This paper reports the first cohort-scale evaluation of this architecture. The evaluation uses the Parkinson’s Progression Markers Initiative (PPMI; [13, 14]; *N*=4,628; 28,185 visits; five-fold cross-validation) and is structured around four questions. First, does the Bayesian architecture produce calibrated distributional forecasts? Second, does the confidence gate yield governed reporting that is measurable, auditable, and equitable? Third, which architectural components are data-informed versus prior-dominated? Fourth, where does the framework identify its own operating boundaries, and what engineering specifications do those boundaries produce? Together, these four questions define the evaluation framework that a governed clinical twin must satisfy. Independent model refits, coupling-topology sensitivity analyses (assessing whether alternative cross-domain influence structures change model conclusions), and extensions for partial observation and MoCA ceiling censoring are included. All findings are within-PPMI. Prospective validation on the National Centre of Excellence in Research on Parkinson’s Disease (NCER-PD), a Luxembourg-based prospective clinical cohort, under routine clinical conditions is pre-specified as the deployment-readiness test. A research prototype at https://symphony-dt.com/ demonstrates implementability (research use only).

The paper makes three contributions. First, a governed Bayesian architecture that couples monotone progression dynamics with uncertainty-gated reporting. Second, a six-rule reporting framework that operationalises when the system must remain silent. Third, a cohort-scale evaluation covering calibration, component attribution, equity, and governance diagnostics.

## 2 Methods

### 2.1 Design rationale

The governed digital twin is a clinic-updated, uncertainty-gated probabilistic model of Parkinson’s disease progression [10, 28]. It tracks three clinical endpoints: motor function (MDS-UPDRS Part III), cognition (MoCA), and autonomic function (SCOPA-AUT). Four deliberate architectural constraints define its scope and govern all downstream inference, evaluation, and reporting:

i. **Monotone latent progression**. Disease severity in each domain is modelled as a componentwise non-decreasing latent process, consistent with the irreversible staging framework established for neurodegenerative diseases [15, 16, 17]. The model class forbids latent improvement, remission, or disease modification within any component. Short-term improvements in observed clinic scores may still arise through measurement noise, but the latent trajectory is irreversible by construction. The constraint deliberately delimits model expressivity to match the degenerative disease assumption and serves as an identifiability safeguard under sparse clinic sampling.
ii. **Clinic-updated estimation**. State estimation and updating occur exclusively at clinic visit times. The model does not incorporate continuous streams from sensors, wearable devices, or derived kinematic or voice features. The model is a *visit-triggered* digital twin, not a continuous-time monitor.
iii. **Uncertainty by design**. Prediction intervals, threshold-event probabilities, and confidence-gated suppression rules are integral to the model’s output specification. Outputs that fail diagnostic or uncertainty criteria are suppressed rather than reported with caveats, operationalising the principle that silence is preferable to unreliable point estimates.
iv. **Non-causal scope**. The model estimates associational progression dynamics conditional on observed history. It does not estimate treatment effects, medication responsiveness, or disease modification. Levodopa-equivalent daily dose (LEDD) is exported for stratified performance reporting only and does not enter the likelihood or latent dynamics.

These constraints are maintained throughout: every modelling choice, inference procedure, evaluation metric, and reporting construct is designed to be consistent with them. Sections 2.2–8 specify the model in full.

### 2.2 Data source and cohort

We used longitudinal observational data from the Parkinson’s Progression Markers Initiative (PPMI), a multicentre prospective observational study designed to identify markers of PD progression [13, 14]. PPMI enrols participants across four primary cohorts: Parkinson’s Disease (confirmed PD), Prodromal (individuals at elevated PD risk), Healthy Control, and SWEDD (Scans Without Evidence of Dopaminergic Deficit). The present analysis uses data from the 28 January 2026 export.

Because the model imposes componentwise non-decreasing latent progression (Section 3.2), which is structurally incompatible with non-degenerative trajectories, we defined a conservative degenerative-trajectory subset for all model fitting and evaluation. Participants were selected using the PPMI cohort designation field (COHORT_DEFINITION), retaining the Parkinson’s Disease and Prodromal cohorts and excluding Healthy Controls (*n* = 400) and SWEDD (*n* = 81). Inclusion was verified by cross-referencing cohort designation labels against the APPRDX code where available (Supplementary Methods Section S1.2). Individual eligibility required non-missing sex and birth date to enable computation of baseline covariates **d**_*i*_, and at least one clinic visit in the master visit index (Supplementary Methods Section S1.4).

The analysis dataset comprises *N* = 4,628 participants (1,829 Parkinson’s Disease; 2,799 Prodromal) and 28,185 clinic visit rows. A clinic visit is defined as a unique (participant, date) pair in the union of outcome assessment dates across the three endpoints; if multiple outcome forms were completed on the same calendar date, the earliest assessment time was used as the canonical visit date. Baseline (*t*_*i*,0_ = 0) is defined as each participant’s first clinic visit date.

Cohort demographic characteristics are reported in Supplementary Table S5.

### 2.3 Preprocessing and outcome derivation

PPMI visit dates are recorded at month resolution (MM/YYYY). All temporal computations anchor to day 15 of the recorded month, applied uniformly to visit dates, birth dates, and medication start dates (Supplementary Table S2). Sensitivity to this convention was assessed by verifying that inter-visit intervals are negligibly affected: the ±14 day jitter is below 2% of the minimum six-month horizon. Time since baseline *t*_*i,n*_ is measured in days, where *t*_*i,n*_ = (visit date_*n*_ − baseline date_*i*_) in days. Age at baseline is computed as age_*i*_ = (baseline date_*i*_ − birth date_*i*_)*/*365.25 in years.

MDS-UPDRS Part III total [30] 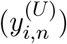, the primary measure of motor severity, was extracted from the PPMI UPDRS export. The raw export contains paired on-/off-medication assessments at some visits (34.4% of raw rows); duplicate (participant, visit) records were resolved deterministically by preferring the row with non-missing total score and breaking ties by earliest assessment time. The resolution strategy selects by assessment completeness rather than medication state, because the model represents the recorded clinical phenotype at each visit rather than an idealised medication-free state; this ensures consistency with routine clinical documentation where both on- and off-medication assessments may be recorded. Values outside [0, 132] were set to missing. MoCA total [31] 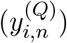, the primary measure of cognitive function, was extracted with support [0, 30]. SCOPA-AUT total [32] 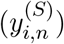, the primary measure of autonomic dysfunction, was derived by summing 25 items, each scored on {0, 1, 2, 3}; a valid total required at least 20 of 25 non-missing items, yielding support [0, 69] (observed maximum in PPMI: 55). No outcome imputation was performed; missing values are encoded with explicit binary flags and treated as missing at random (MAR) conditional on observed history for all inference.

LEDD was harmonised from the PPMI Concomitant Medication Log following the official PPMI LEDD methodology [29]. Per-visit LEDD is the sum of LEDD values for all medication records active at the visit month, with missing stop dates treated as ongoing. Consistent with the non-causal scope boundary (Section 2.1), LEDD does *not* enter the likelihood or latent dynamics; it is used exclusively for stratified descriptive evaluation summaries.

Genetic consensus data (assay-availability flags and PD-gene variant annotations) are available for 3,345 participants (72.3%). Only draws with draw time 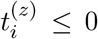 (days relative to baseline) are eligible; among eligible draws, the latest pre-baseline draw is selected. Participants with no pre-baseline draw are assigned **z**_*i*_ = **0** and marked omics-missing for reporting. Raw features are exported without standardisation or pathway scoring; both operations are performed within training folds at model-fitting time to prevent leakage (Section 3.4).

Complete preprocessing details, including stepwise attrition counts, duplicate-resolution audit tables, outcome distributional summaries, omics feature coverage, and leakage-control specifications, are provided in the Supplementary Methods.

### 2.4 Notation and exported variables

Participants are indexed by *i* ∈ {1, …, *N*}. Clinic visit times for participant *i* are 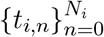 in days since baseline *t*_*i*,0_ = 0, with *t*_*i,n*+1_ *> t*_*i,n*_. Define Δ*t*_*i,n*_ = *t*_*i,n*+1_ − *t*_*i,n*_.

Disease status is represented by a three-dimensional latent state

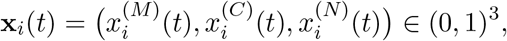

where superscripts (*M, C, N*) denote motor, cognitive, and global non-motor severity (larger values indicate worse severity). We introduce an unconstrained latent process **s**_*i*_(*t*) ∈ ℝ^3^ with **x**_*i*_(*t*) = *σ*(**s**_*i*_(*t*)), where

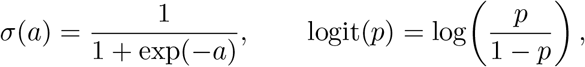

applied elementwise.

Exported clinical outcomes at visit *n* are

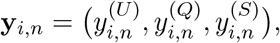

where *y*^(*U*)^ is MDS-UPDRS Part III total, *y*^(*Q*)^ is MoCA total, and *y*^(*S*)^ is SCOPA-AUT total. Score ranges are fixed:

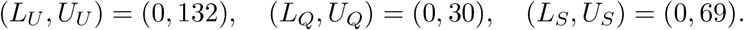

The notation uses two label sets by design. Latent states use disease-domain labels: *M* (motor), *C* (cognitive), *N* (non-motor). Observed outcomes use instrument labels: *U* (UPDRS-III), *Q* (MoCA), *S* (SCOPA-AUT). The one-to-one mapping is *M* → *U, C* → *Q, N* → *S*. Coupling parameters (Section 3.2) use the disease-domain labels (*c*_*MM*_, *c*_*CM*_, etc.) because they describe latent-state interactions, not instrument-level relationships.

Baseline covariates are **d**_*i*_ = (age_*i*_, 𝕀{sex_*i*_ = male})^⊤^, where

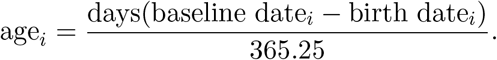

Sex and birth date are required; participants missing either are excluded. Omics-derived pathway scores are **z**_*i*_ ∈ ℝ^*P*^ (Section 3.4); if omics are unavailable, **z**_*i*_ = **0**.

#### Missingness and scope

Let 𝒦_*i,n*_ ⊆ {*U, Q, S*} denote the set of observed outcomes at visit (*i, n*). Missing outcomes are handled by omitting the corresponding likelihood factor (Section 3.3). No explicit MNAR or dropout model is introduced; missingness is treated as MAR conditional on observed history, with sensitivity analyses prespecified in Section 6.4. The model requires only exported clinic visit times, the three clinic totals, sex, birth date, and (optionally) pre-baseline omics draws. No raw audio/video, sensor streams, or derived kinematic/voice features are used.

Table 1 provides a consolidated reference for all notation used in the model specification.

**Table 1:** Notation reference. All symbols are defined at first use in the text; this table serves as a consolidated index.

| Symbol | Domain | Definition |
| --- | --- | --- |
| $\mathbf{x}_i(t)$ | $(0, 1)^3$ | Latent disease state (motor, cognitive, non-motor) |
| $\mathbf{s}_i(t)$ | $\mathbb{R}^3$ | Unconstrained latent process; $\mathbf{x}_i(t) = \sigma(\mathbf{s}_i(t))$ |
| $\mathbf{y}_{i,n}$ | $\mathbb{R}^3$ | Observed clinical outcomes (UPDRS-III, MoCA, SCOPA-AUT) |
| $\mathbf{r}_i$ | $\mathbb{R}^3$ | Participant-specific progression drive |
| $\mathbf{C}$ | $\mathbb{R}^{3 \times 3}$ | Cross-domain coupling matrix (sparse; Section 3.2) |
| $\boldsymbol{\xi}_i(t, \Delta t)$ | $(0, \infty)^3$ | Multiplicative log-normal innovation |
| $\boldsymbol{\sigma}_\xi$ | $[0, \infty)^3$ | Process-variability scales |
| $\mathbf{d}_i$ | $\mathbb{R}^2$ | Baseline covariates (age, sex indicator) |
| $\mathbf{z}_i$ | $\mathbb{R}^P$ | Omics-derived pathway scores |
| $\boldsymbol{\Gamma}$ | $\mathbb{R}^{3 \times 2}$ | Demographic effect on progression drive |
| $\mathbf{A}$ | $\mathbb{R}^{3 \times P}$ | Pathway effect on progression drive |
| $h(p; a, b)$ | $(0, 1)$ | Monotone nonlinear observation link |
| $\sigma_k$ | $\mathbb{R}_{>0}$ | Observation noise SD for endpoint $k$ |
| $\Phi$ | — | Full set of global parameters (Section 3.5) |
| $\mathcal{D}_{i, \leq t_{i,n}}$ | — | Information set at visit $t_{i,n}$ |
| $\Delta_{i,n}$ | $\mathbb{R}_{\geq 0}$ | Per-visit latent uncertainty (max IQR; Section 5.4) |
| $\tau_{\text{conf}}$ | $\mathbb{R}_{>0}$ | Confidence-gating threshold (Section 5.4) |

## 3 Clinic-updated digital twin: model specification

### 3.1 Latent state constraint

Disease status is represented by **x**_*i*_(*t*) ∈ (0, 1)^3^ with **x**_*i*_(*t*) = *σ*(**s**_*i*_(*t*)). The latent dynamics impose *componentwise non-decreasing progression* of **x**_*i*_(*t*) over time: the model class forbids latent improvement, remission-like trajectories, or disease modification within a component. Short-term improvements in observed clinic scores may still occur through the observation noise model (Section 3.3), but the latent process remains non-decreasing by construction. This constraint is the primary identifiability safeguard of the model and reflects the degenerative disease assumption that motivates cohort selection (Section 2.2).

### 3.2 Monotone latent dynamics with sparse cross-domain coupling

A transition is defined for any Δ*t >* 0, which fully specifies forecasting as forward simulation under the model. For any *t* and Δ*t >* 0,

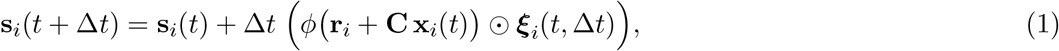

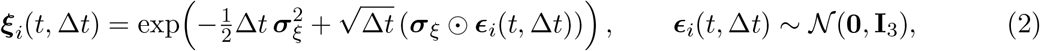

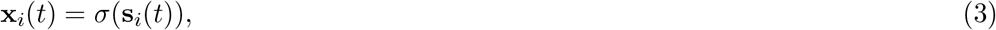

where *ϕ*(*a*) = log(1 + exp(*a*)) is applied elementwise, ⊙ is elementwise multiplication, and

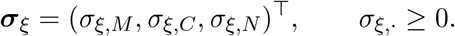

This parameterisation satisfies 𝔼[*ξ*_*i,j*_(*t*, Δ*t*)] = 1 for each dimension *j*, yielding the conditional mean increment

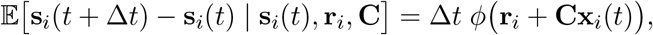

and enforces non-negative increments componentwise because *ϕ*(·) ≥ 0 and ***ξ***_*i*_(*t*, Δ*t*) *>* **0** componentwise.

The participant-specific progression drive **r**_*i*_ ∈ ℝ^3^ encodes each individual’s baseline rate of disease advancement across the three domains. Cross-domain coupling is encoded by **C** ∈ ℝ^3*×*3^ with fixed sparse topology

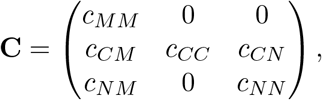

which prevents cyclic feedback and limits degrees of freedom. The off-diagonal entries encode the assumption that motor impairment may influence cognitive and autonomic progression, consistent with predominant staging models in which motor deficit precedes or drives multi-domain spread [12]. Bidirectional coupling is biologically plausible; autonomic dysfunction can precede motor symptoms in prodromal PD [35]. However, under PPMI’s sparse clinic sampling, a fully connected 3 × 3 matrix would introduce nine free parameters unlikely to be jointly identifiable. The chosen topology restricts coupling to six parameters whose identifiability is verified empirically via contraction ratios (Section 6.4); alternative topologies can be tested with richer longitudinal data. The diagonal entries *c*_*MM*_, *c*_*CC*_, *c*_*NN*_ represent state-dependent acceleration: the effective drive 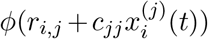 becomes state-dependent, capturing the empirical observation that progression tends to accelerate with advancing severity. These are mechanistically distinct from the participant-specific drives **r**_*i*_, which are constant baseline rates.

#### Sampled objects versus induced trajectories

The objects directly sampled in inference are (**s**_*i*_(*t*_*i*,0_), **r**_*i*_), global parameters **Φ**, and the innovation variables {***ϵ***_*i*_(*t*, Δ*t*)} (equivalently {***ξ***_*i*_(*t*, Δ*t*)}). The innovations {***ϵ***_*i*_(*t*_*i,n*_, Δ*t*_*i,n*_)} are included as explicit latent variables in the inference target density; conditional on these and the initial state, the visit-time trajectory is deterministic. The latent trajectory {**s**_*i*_(*t*)} at any collection of times is not separately parameterised; it is deterministically induced by recursion through Eq. (1) given the sampled initial state, parameters, and innovations.

#### Deterministic construction on observed visit times

For fixed (**s**_*i*_(*t*_*i*,0_), **r**_*i*_, **Φ**) and sampled 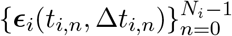, the visit-time trajectory 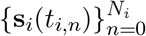 is constructed deterministically by recursion using Eq. (1). Forecasting to any future time is defined analogously as forward simulation using newly sampled innovations over the forecast increment, conditional on the posterior at the conditioning time.

### 3.3 Truncated-Gaussian observation model

Each endpoint *k* ∈ {*U, Q, S*} is modelled as truncated Gaussian:

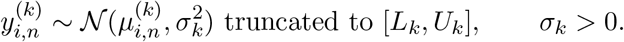

Expected scores use a monotone nonlinear link. Define

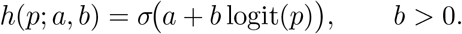

Then

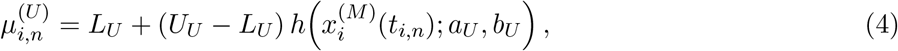

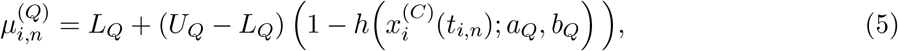

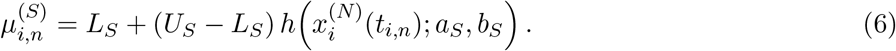

Thus UPDRS-III and SCOPA-AUT increase with severity while MoCA decreases with severity. LEDD does not enter 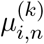; the model does not represent symptom modulation by medication and does not support causal medication-effect statements.

The truncated-Gaussian specification accommodates the non-trivial boundary mass observed in the data: 14.2% of MoCA values lie at ceiling (score 30) and 10.3% of UPDRS-III values lie at floor (score 0); see Supplementary Methods Section S1.8. Boundary observations are handled jointly by the measurement model (truncated likelihood concentrates probability mass near the boundary) and by reporting policy (threshold-event probabilities are suppressed at boundary values; Section 5.4, rule 5). A formal censoring model with explicit point mass at boundaries is a planned extension for populations with higher boundary prevalence.

### 3.4 Omics pathway scores as prior covariates

The architecture accommodates optional omics-derived pathway scores **z**_*i*_ ∈ ℝ^*P*^ as baseline covariates shifting the prior mean of **r**_*i*_ (Eq. 7), consistent with the small marginal genetic effects observed in prior PPMI analyses [19]. Only pre-baseline draws are used; participants without a pre-baseline draw are assigned **z**_*i*_ = **0**. Pathway definitions are derived from established PD-relevant gene sets (*P* ≤ 10; Supplementary Table S9) and are fixed before model fitting. Feature standardization and pathway scoring are performed within training folds to prevent leakage. Full specification of pathway construction, standardisation, and the identifiability cap is provided in Supplementary Section S6.

### 3.5 Hierarchical structure, priors, and identifiability safeguards

Participant progression drives are partially pooled [18]:

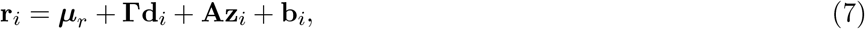

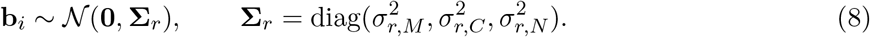

This structure provides partial pooling under sparse visits and makes explicit which components are learned from repeated clinical outcomes versus regularised by prior structure. In particular, the coupling parameters in **C**, the process-variability scales ***σ***_*ξ*_, and pathway effects **A** are weakly identified in typical sparse clinic follow-up and are therefore shrinkage-regularised by design; posterior contraction and stability diagnostics are used to assess whether these components are data-informed (Section 6.4).

Global parameters are

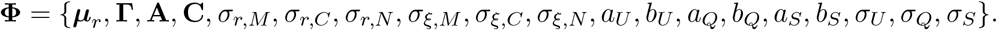

Priors are:

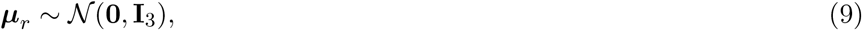

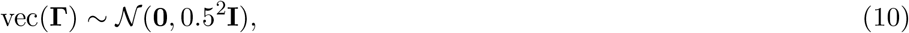

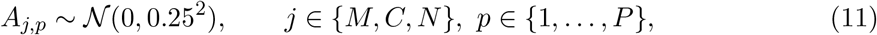

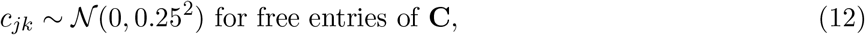

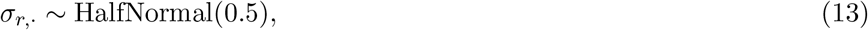

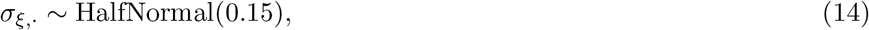

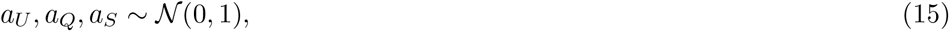

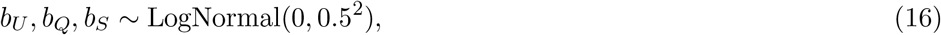

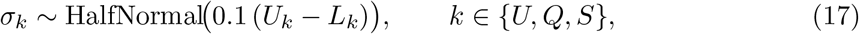

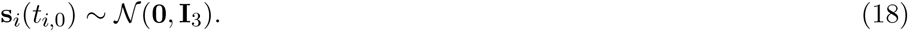

Prior scales are weakly informative and chosen to encode the following expectations: *σ*_*ξ*,·_ ~ HalfNormal(0.15) centres process variability near zero, regularising toward smooth progression consistent with the expectation that visit-to-visit latent change is small relative to measurement noise; *A*_*j,p*_ ~ 𝒩 (0, 0.25^2^) enforces strong shrinkage on pathway effects, reflecting the a priori expectation that individual genetic features have small marginal effects on progression rate; *c*_*jk*_ ~ 𝒩 (0, 0.25^2^) applies equivalent shrinkage to coupling, limiting cross-domain influence to be data-driven rather than prior-driven. Sensitivity to prior scales is assessed via contraction ratios (Section 6.4).

## 4 Inference, updating, and uncertainty-aware forecasting

### 4.1 Cross-validated training protocol

All inference and evaluation are performed under 5-fold cross-validation with participant-level splits stratified by cohort designation (Parkinson’s Disease vs. Prodromal). This protocol evaluates generalisation to unseen participants under the clinic-updated setting. Within each fold:

- omics standardisation statistics (Section 3.4) are computed on the training fold only;
- bootstrap stability for pathway effects (Section 5.3) is computed on the training fold only;
- confidence-gating thresholds (Section 5.4) are computed on the training fold only;
- the adaptive global-uncertainty subsample size *L*_cond_ (Section 4.3) is selected on the training fold only;
- model fitting is performed on the training fold and evaluated on the held-out fold without refitting.

### 4.2 Population-level posterior inference (offline)

Within each fold, we sample from the posterior over **Φ**, participant parameters (**s**_*i*_(*t*_*i*,0_), **r**_*i*_) for *i* ∈ ℐ_train_, and the innovation variables {***ϵ***_*i*_(*t*_*i,n*_, Δ*t*_*i,n*_)} implied by Eq. (2) using Hamiltonian Monte Carlo with the No-U-Turn Sampler (NUTS) [22], implemented in Stan [36]. The innovations are included as explicit latent variables in the NUTS target density; conditional on these and the initial state, visit-time trajectories are deterministically induced by recursion through Eq. (1). We run 4 chains with 1,000 warm-up and 1,000 sampling iterations per chain; the *L* = 200 posterior draws used for downstream propagation are obtained by thinning the concatenated post-warm-up draws at uniform spacing across chains, reducing downstream propagation cost while preserving a representative subset of the posterior.

Convergence requirements are: 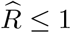 for all scalar components of **Φ** and selected trajectory summaries (posterior means of **x**_*i*_(*t*_*i,n*_)), bulk effective sample size ≥ 400 for these quantities, and zero divergent transitions after warm-up.

### 4.3 Global-uncertainty propagation with adaptive subsampling

Let 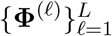 be *L* = 200 posterior draws from the training-fold posterior (Section 4.2). To reduce computation while preserving uncertainty propagation, we choose a fold-specific subsample size *L*_cond_ from the candidate set

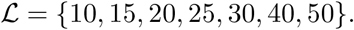

For each candidate *L*_cond_ ∈ ℒ, draw a subsample uniformly without replacement from {1, …, *L*} and compute one-step-ahead predictive means and 95% interval widths for a fixed set of 50 training participants (selected once per fold by a fixed random seed; 50 participants provide sufficient statistical power to detect clinically relevant changes in predictive means, with *<*5% false negative rate for differences exceeding 0.5 UPDRS-III points based on pilot variance estimates). Choose the smallest *L*_cond_ such that, relative to the next larger candidate in ℒ, the mean absolute change in predictive means is ≤ 0.5 for UPDRS-III, ≤ 0.2 for MoCA, and ≤ 1.0 for SCOPA-AUT (corresponding to approximately 0.4%, 0.7%, and 1.4% of the respective endpoint ranges, well below established minimal clinically important differences: 2.3–2.7 points for UPDRS-III [20, 21] and approximately 1–2 points for MoCA), and the mean relative change in 95% interval width is ≤ 10% for all three endpoints. The selected *L*_cond_ and the corresponding subsample indices are fixed for the fold and reused for all participants and all evaluation metrics. All reported uncertainty intervals and calibration assessments correspond to this explicit global-uncertainty propagation scheme.

### 4.4 Clinic-updated personalisation and sequential conditioning

For participant *i*, define the information set at visit time *t*_*i,n*_:

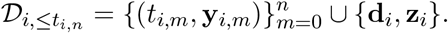

For each *ℓ* in the fold-specific subsample of size *L*_cond_, we sample from the participant-conditional posterior

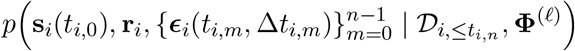

using NUTS. Sequential conditioning is performed at each new clinic visit by warm-starting the sampler from the previous visit’s final state and introducing the new innovation variable for the interval (*t*_*i,n*_, *t*_*i,n*+1_]. The global parameters **Φ**^(*ℓ*)^ are held fixed; only participant-level quantities are updated. This conditioning scheme is clinic-triggered and does not assume continuous real-time coupling.

#### Computational budget and failure handling

For each (*i, n, ℓ*) we cap warm-up at 250 iterations and sampling at 250 iterations per chain with 4 chains. If any of the following occurs: (i) divergent transitions after warm-up; (ii) mean acceptance probability outside [0.7, 0.9]; (iii) 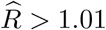 for any component of **r**_*i*_, then the update at (*i, n*) is marked as failed for that *ℓ* and excluded from the mixture; if fewer than max{10, ⌈0.4*L*_cond_⌉} conditional runs pass diagnostics at (*i, n*), all forecasts and threshold probabilities for that conditioning visit are suppressed (Section 5.4). This operationalises uncertainty-aware gating at the inference level rather than reporting point estimates by default.

### 4.5 Uncertainty-propagating forecasting by forward simulation

Forecasting is defined as forward simulation under the constrained latent process, conditional on the posterior at the conditioning visit. Forecasts are produced only for (i) the next observed visit time *t*_*i,n*+1_ (one-step-ahead) and (ii) horizon-binned targets within ±30 days of ℋ = {180, 365, 730} days (Section 6.2). For each forecast instance with target time *t*^⋆^ *> t*_*i,n*_, set Δ*t*^⋆^ = *t*^⋆^ − *t*_*i,n*_ and generate *S* = 2,000 posterior predictive draws by repeating the following procedure, distributing draws across the *L*_cond_ successful conditional runs (approximately *S/L*_cond_ draws per global parameter setting):

1. sample *ℓ* uniformly from the set of successful conditional runs at (*i, n*);
2. draw (**s**_*i*_(*t*_*i*,0_), **r**_*i*_) and the past innovations from the participant-conditional posterior under **Φ**^(*ℓ*)^;
3. induce **s**_*i*_(*t*_*i,n*_) deterministically by recursion through Eq. (1) on observed intervals up to *t*_*i,n*_;
4. propagate from *t*_*i,n*_ to *t*^⋆^ using Eq. (1) with increment Δ*t*^⋆^ and a fresh innovation draw ***ϵ***_*i*_(*t*_*i,n*_, Δ*t*^⋆^) ~ 𝒩 (**0, I**_3_);
5. map **x**_*i*_(*t*^⋆^) = *σ*(**s**_*i*_(*t*^⋆^)) to endpoint means and sample outcomes from the truncated Gaussian likelihoods.

This procedure propagates parameter uncertainty (through **Φ**^(*ℓ*)^), patient-level uncertainty (through the conditional posterior), process uncertainty (through innovations), and measurement uncertainty (through *σ*_*k*_). Forecasts do not represent latent improvement or medication-driven disease modification, because the latent dynamics are monotone by construction and medication does not enter the generative model.

### 4.6 Posterior predictive checks

Within each training fold, posterior predictive checks (PPCs) are computed from posterior draws and assessed using fixed discrepancy measures [33]:

#### Residual calibration under truncation (PIT)

For each observed (*i, n, k*), compute the posterior mean parameters 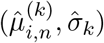 and the truncated-normal CDF 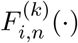 on [*L*_*k*_, *U*_*k*_]. Define

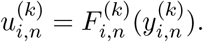

Compute the Kolmogorov–Smirnov distance between the empirical distribution of 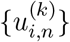 and Uniform(0, 1). Flag endpoint *k* in a fold if KS *>* 0.10. To detect truncation-induced miscalibration not visible in the aggregate KS statistic, PIT calibration is additionally reported stratified by boundary proximity (observations within 1 unit of *L*_*k*_ or *U*_*k*_).

#### Increment distribution discrepancy

For each endpoint *k*, compute empirical increments 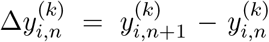 and replicated increments from posterior predictive draws at the same Δ*t*_*i,n*_. For each quartile bin of Δ*t*_*i,n*_, compute absolute differences in the 10th, 50th, and 90^th^ percentiles between observed and replicated increments; flag a bin if any absolute difference exceeds 0.1(*U*_*k*_ − *L*_*k*_).

#### Cross-endpoint residual dependence

Using truncated-normal residuals 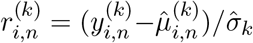, compute the empirical residual correlation matrix across (*U, Q, S*) and compare to the posterior predictive distribution of residual correlations. Flag a fold if the observed Frobenius norm distance to the posterior predictive mean exceeds the 95% posterior predictive quantile of that distance.

## 5 Intrinsic explainability and reporting constraints

### 5.1 Scope of intrinsic explainability

Explainability is defined as intrinsic, parameter-based explainability of the probabilistic generative model. Explanations are computed as functionals of posterior distributions over model parameters and latent states, not as post-hoc attributions for a black-box predictor. Because the twin is clinic-updated and excludes sensor streams, no feature-attribution explanations for high-frequency signals are defined. Explanations are interpreted as model-implied contributions to monotone progression under the constrained model class and are not construed as causal mechanisms.

### 5.2 Mechanistic parameter explanations

Let *π*_*i,n*_ denote the posterior at visit time *t*_*i,n*_ under the fold-specific mixture. We report posterior summaries for:

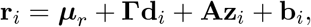

including each additive component per dimension *j* ∈ {*M, C, N*}, with **b**_*i*_ computed deterministically per draw. We also report the pre-softplus increment predictor

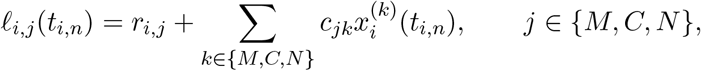

and its decomposition into *r*_*i,j*_ and (**Cx**_*i*_(*t*_*i,n*_))_*j*_.

### 5.3 Stability gates and explanation policy

For any scalar explanation quantity *T*, compute sign probability Pr(*T >* 0 | 𝒟_*i*,≤*t*_); explanations are suppressed unless this exceeds 0.95.

For participants with *N*_*i*_ ≥ 3, local explanations are additionally required to satisfy leave-one-visit-out stability: delete each interior visit once, recompute the participant posterior under the same fold-specific mixture, and suppress an explanation if the maximum absolute change in posterior mean exceeds 0.25 on the *ℓ*-scale or if the minimum sign probability across deletions is below 0.90.

Population pathway effects *A*_*j,p*_ are subject to bootstrap sign stability (*B* = 200 resamples; threshold ≥ 0.90). This policy reflects the weak identifiability of pathway effects under sparse clinic sampling (Supplementary Section S6).

### 5.4 Confidence gating and output suppression

Define per-visit latent uncertainty

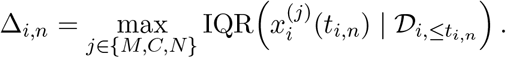

Within each training fold, choose *τ*_conf_ ∈ {0.05, 0.06, …, 0.60} to 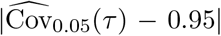 subject to 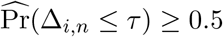, where 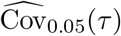 is one-step-ahead 95% interval coverage restricted to anchors with Δ_*i,n*_ ≤ *τ* on the training fold. The threshold is selected on training-fold data only; all reported coverage metrics are on held-out test-fold participants. The narrow cross-fold range of *τ*_conf_ (0.17–0.19) emerged empirically from the training-fold grid search and was not pre-specified, confirming calibration stability across data splits.

Outputs are suppressed if any of the following hold at the conditioning visit:

1. fewer than two clinic visits exist for the participant;
2. any of 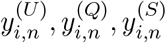 is missing at the conditioning visit (a conservatism constraint for initial deployment; the observation model accommodates partial missingness via likelihood-factor omission, but conditioning on incomplete visit data is deferred to future evaluation);
3. Δ_*i,n*_ *> τ*_conf_;
4. fewer than max{10, ⌈0.4*L*_cond_⌉} conditional runs pass diagnostics at (*i, n*);
5. any conditioning-visit outcome equals its boundary 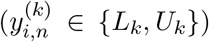, in which case only predictive intervals are shown and no threshold-event probabilities are reported;
6. LEDD at the anchor visit exceeds 500 mg for UPDRS-III outputs (medication-context rule). Because LEDD does not enter the observation model, motor-score variability under high medication burden is unmodelled, producing systematic under-dispersion (Section 9.7). Anchor-visit LEDD is known at forecast time, making this rule operationally feasible in prospective deployment. This rule suppresses UPDRS-III forecasts where calibration is known to degrade.

Rules 1–5 are structural gating conditions derived from the model architecture. Rule 6 is a context-aware extension that addresses a known scope limitation. Additional context rules may be added as validation identifies further calibration boundaries, provided each rule is pre-specified, evaluated, and documented before deployment. These rules enforce that uncertainty is propagated, evaluated, and gated rather than treated as an optional annotation.

## 6 Evaluation and validation

### 6.1 Splits, leakage control, and evaluation scope

We use 5-fold cross-validation with participant-level splits stratified by cohort designation (Section 4.1). All fold-dependent quantities (omics standardisation, bootstrap stability, *τ*_conf_, and the adaptive *L*_cond_ selection) are computed on the training fold only [24]. Forecast evaluation uses rolling-origin conditioning on 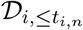. Evaluation uses only clinic-exported variables and optional pre-baseline omics pathway scores; LEDD is used only for stratified reporting and is not interpreted as a causal driver.

### 6.2 Forecast tasks under the monotone latent process

#### One-step-ahead

For each anchor (*i, n*) with *n < N*_*i*_, evaluate the posterior predictive distribution of each endpoint observed at *t*_*i,n*+1_.

#### Horizon-binned forecasting under irregular follow-up

Define horizon centres ℋ = {180, 365, 730} and tolerance *δ*_*h*_ = 30. For each anchor (*i, n*) and each bin *h*, evaluate targets *m > n* with |(*t*_*i,m*_ − *t*_*i,n*_) − *h*| ≤ *δ*_*h*_. Predictions are generated with Δ*t* = *t*_*i,m*_ − *t*_*i,n*_ using Eq. (1). Metrics are reported by bin and stratified by realised Δ*t* quartiles to detect visit-density artefacts. Horizon-binned evaluation conditions on visit existence at the target time; participants who drop out before reaching a horizon bin are not evaluated for that bin. Because dropout may be severity-related, metrics at longer horizons may reflect a healthier-surviving subpopulation; the follow-up count stratification in Section 6.4 is designed to quantify this selection effect.

### 6.3 Metrics and uncertainty evaluation

For each forecast instance, generate *S* = 2,000 posterior predictive draws under the fold-specific mixture (Section 4.5). Let 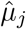 be the predictive mean. To avoid domination by dense follow-up, compute participant-level averages first. For MAE,

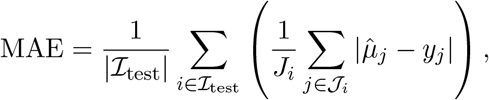

with analogous participant-level RMSE. Distributional accuracy is assessed by CRPS estimated from draws and aggregated by participant-level averaging:

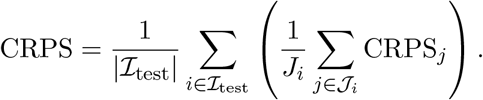

Calibration is assessed via 95% predictive interval coverage and threshold-event calibration (Brier score and reliability curves) for:

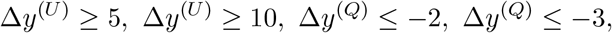

computed relative to the anchor visit. Threshold-event calibration is computed only for instances where the endpoint is observed at both anchor and target visits and where the anchor outcome is not at a boundary value (Section 5.4, rule 5); missing anchor or target values are excluded. Reported calibration corresponds to the same uncertainty propagation regime used in forecasting and gating (Sections 4.3–5.4).

### 6.4 Ablations, robustness, and identifiability diagnostics

Ablations compare: (i) full model, (ii) no coupling (off-diagonals of **C** set to zero), (iii) no omics (**A** = **0**), (iv) linear-link variant (replace *h*(*p*; *a, b*) by *h*(*p*) = *p*). Each variant is refit and evaluated under the identical protocol and the same global-uncertainty propagation scheme.

Robustness is assessed by stratifying metrics by baseline age quartile and baseline LEDD quartile (LEDD only for stratification, not for causal inference). Suppression rates (Section 5.4) are reported stratified by baseline outcome severity quartile to verify that gating does not preferentially suppress outputs for high-severity participants. Missingness sensitivity is assessed by reporting metrics by follow-up count strata and comparing baseline posterior means across strata.

#### Posterior contraction reporting

To quantify whether coupling and pathway effects are data-informed rather than prior-dominated, report contraction ratios on the training fold:

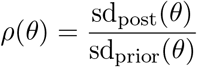

for each free coupling parameter in **C** and each pathway effect *A*_*j,p*_. Posterior SDs are computed from NUTS draws; prior SDs are the prior scales (0.25 for both coupling and *A*_*j,p*_). Ratios *ρ*(*θ*) *<* 0.5 indicate meaningful posterior contraction beyond the prior; ratios near 1.0 indicate the parameter is prior-dominated and should not be interpreted as data-driven. Ratios are summarised by median and interquartile range across folds, alongside posterior sign probabilities Pr(*θ >* 0 | 𝒟_train_).

### 6.5 Computational feasibility reporting

Including every innovation ***ϵ***_*i*_(*t*_*i,n*_, Δ*t*_*i,n*_) as an explicit latent variable in NUTS scales the target density dimension with the total number of inter-visit intervals (≈13,000 in the training fold). The sequential conditioning step further multiplies computation by up to *L*_cond_ conditional runs per participant per visit. To ensure that the inference scheme is computationally tractable at this scale, we report the following for each fold: (i) wall-clock time for population-level posterior sampling and for the full sequential-conditioning pass, reported per chain and as total elapsed time; (ii) effective sample sizes (ESS) summarised by parameter class: global parameters **Φ**, participant-level drives **r**_*i*_, and innovation variables ***ϵ***_*i*_; (iii) the selected *L*_cond_ value and subsample indices; (iv) the fraction of conditional runs that pass NUTS diagnostics (no post-warm-up divergences, acceptance probability in [0.7, 0.9], *R* ≤ 1.01) across all (*i, n, ℓ*); and (v) the fraction of visits where forecasts are suppressed due to the “fewer than max{10, ⌈0.4*L*_cond_⌉} runs passed” rule. These quantities are computed on the test fold only for deployment-faithful assessment.

### 6.6 Architectural component assessment

To assess the contribution of each architectural component, simplified model variants were fitted on training data and evaluated on held-out test data across all five folds. Two empirical Bayes LME variants were fitted: one with the nonlinear observation link applied to scores before fitting, and one on raw (linearly mapped) scores. Both used per-participant random intercept and slope with partial pooling. This constitutes an independent refit that re-estimates all participant-level parameters under each model variant. Coupling contribution was assessed through posterior contraction ratios (Section 6.4) and cross-domain forecast coherence: Pearson correlations between predicted scores across endpoint pairs at governed anchor visits. Omics contribution was assessed through the pathway stability diagnostics (Section 5.3).

### 6.7 Sex-stratified equity analysis

Suppression rates were stratified by sex at the anchor level across all five folds. Sex was linked through the fold assignment table (*N* =2,491 male; *N* =2,137 female). Cramér’s *V* quantified the effect size of the sex-suppression association. Suppression decomposition by rule identified which gating rule drives any observed difference. Among governed forecasts, MAE and 95% coverage were reported by sex and endpoint. Participant-level governed coverage (≥1 governed visit) was stratified by sex and cohort.

### 6.8 Calibration by follow-up density

Forecast accuracy was stratified by participant visit count in bins: 2–3, 4–6, 7–10, and *>*10 visits. MAE, 95% coverage, interval width, and mean latent uncertainty (Δ_*i,n*_) were reported per bin. Suppression rates by visit-count bin were reported to assess whether sparser follow-up increases governance barriers.

### 6.9 Coupling topology sensitivity

To assess robustness to the unidirectional coupling assumption, *c*_*MN*_ (autonomic to motor, currently fixed at zero) was analytically estimated across all five folds. Motor latent increments were computed from the posterior latent trajectories. Residuals after the current model’s predicted drift were regressed on the coupling covariate Δ*t* · *σ*(drive_*M*_) · *x*_*N*_. Bayesian linear regression with the same 𝒩 (0, 0.25^2^) prior as all coupling entries produced posterior estimates, contraction ratios, and sign probabilities. The analysis was stratified by motor severity quartile to distinguish coupling from severity-dependent confounding.

### 6.10 Partial-observation and censoring extensions

A 2-of-3 domain partial-observation extension was evaluated by projecting governed coverage under a relaxed Rule 2 that requires only two of three endpoints at the conditioning visit. The marginalisation-induced interval widening was estimated from the coupling matrix and domain-specific latent IQRs. A MoCA censoring model was evaluated by replacing the truncated-Gaussian density at *y*=30 with an explicit point mass *P* (*Y* =30) = *P* (*Y* ^∗^ ≥ 30) and computing corrected PIT values and KS statistics.

## 7 Translational layer: from probabilistic outputs to clinical interpretation

### 7.1 Model outputs and reported constructs

The governed digital twin produces four primary output types, each with explicit scope boundaries and uncertainty quantification:

#### State trajectories and evolution

For each participant *i* with at least two clinic visits, the model outputs posterior distributions over latent states 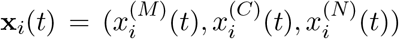 at observed visit times *t*_*i,n*_. These are reported as posterior means and interquartile ranges, reflecting within-participant progression under the monotone constraint. State trajectories are *not* directly observable and do not correspond to any single clinical scale; they represent model-implied disease severity on a (0, 1) continuum for each domain.

#### Outcome forecasts with prediction intervals

At each conditioning visit (*i, n*), the model generates posterior predictive distributions for clinical outcomes at future visits: one-step-ahead (next observed visit *t*_*i,n*+1_) and horizon-binned targets within ±30 days of {180, 365, 730} days. Forecasts are reported as predictive means and 90% or 95% prediction intervals, incorporating parameter uncertainty, patient-level uncertainty, process uncertainty, and measurement uncertainty (Section 4.5). Because the latent dynamics are monotone by construction and medication does not enter the generative model, forecasts do not represent latent improvement or medication-driven disease modification.

#### Clinical event probabilities

The model outputs threshold-crossing probabilities for pre-specified clinical events: 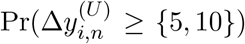 (UPDRS-III worsening), 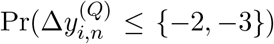 (MoCA decline), computed from posterior predictive draws relative to the conditioning visit. Event probabilities are suppressed when the conditioning-visit outcome equals its boundary (Section 5.4) and are *not* causal effect estimates; they represent model-implied progression risk under the observed covariate profile.

#### Confidence gating and output suppression

Outputs are suppressed according to the rules in Section 5.4. Suppression is reported explicitly rather than filled with default values, operationalising uncertainty-aware gating.

### 7.2 Dashboard and reporting constructs

The clinical dashboard and automated reports map probabilistic outputs to interpretable constructs without introducing causal language. Table 2 defines each reported construct. Key principles:

- **Risk curves and volatility indices** are derived from posterior predictive quantiles and spread metrics, not from hazard models or survival analysis.
- **State occupancy summaries** use discretised latent state bins (e.g., *x*^(*M*)^ ∈ [0, 0.33) labelled “low motor severity”) for interpretability; bin boundaries are fixed a priori and do not adapt to data.
- **Trajectory clusters** are assigned via deterministic rules on observed outcome progressions, not via latent clustering of individual-level parameters **r**_*i*_. Cluster labels (e.g., “stable”, “accelerating”) describe observed patterns, not predicted futures.
- **LEDD stratification** is descriptive only. LEDD does not enter the likelihood or latent dynamics; stratified performance summaries do not estimate medication effects or support causal inference about treatment responsiveness.

**Table 2:**
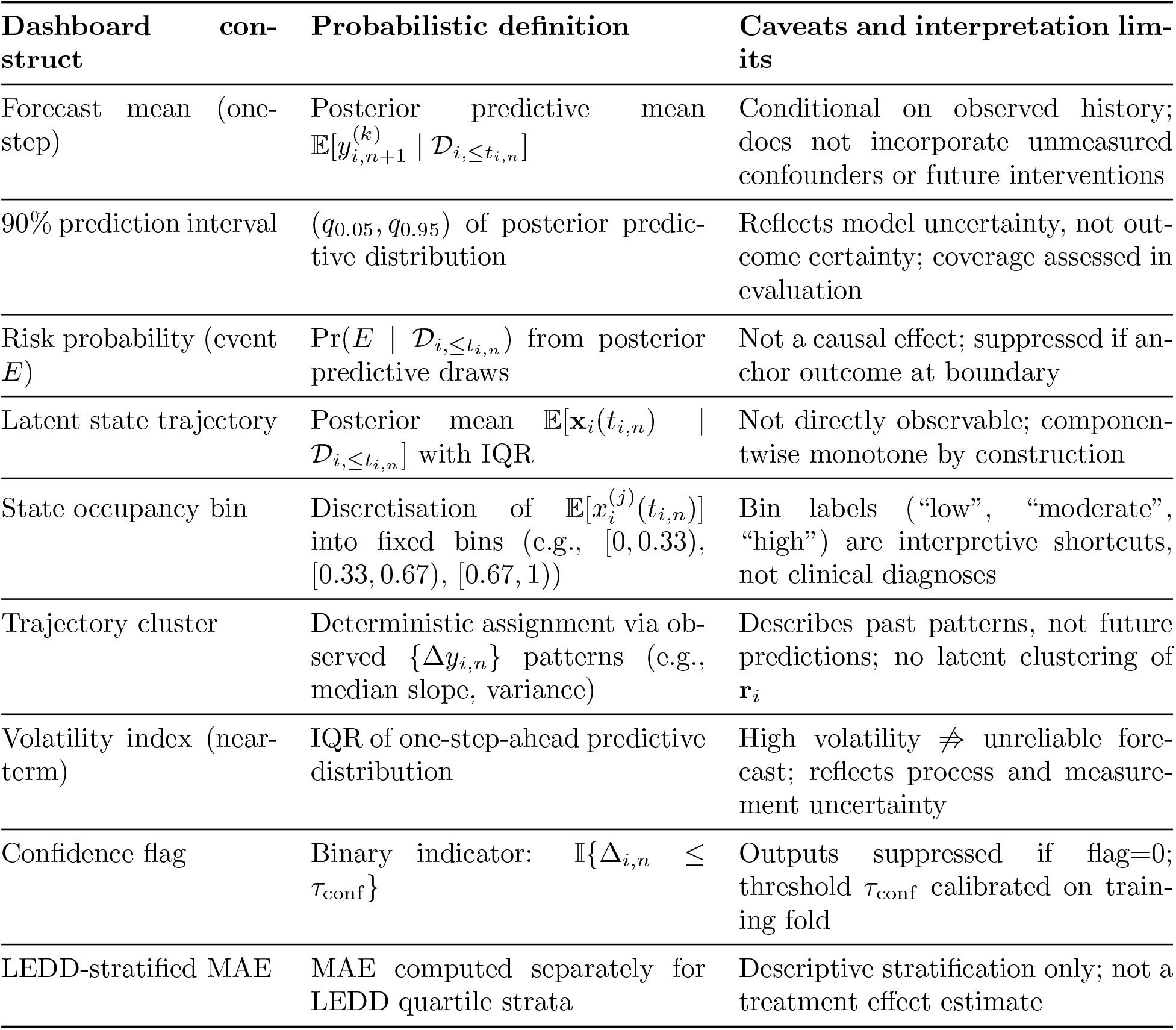
Mapping from model outputs to clinical dashboard constructs. All constructs are derived from posterior predictive distributions or deterministic rules on observed data; none involve causal estimation.

### 7.3 Explicit scope boundaries

The following are *out of scope* and are not supported by the model or reported outputs:

- **Medication causality**: LEDD is exported for stratified reporting only. The model does not identify medication responsiveness, treatment effects, or disease modification. Any medication-related summaries are descriptive performance stratifications.
- **Remission or latent improvement**: The latent process is monotone by construction. Short-term improvements in observed scores may occur via measurement noise, but the latent trajectory **x**_*i*_(*t*) is componentwise non-decreasing. The model class cannot represent remission-like trajectories.
- **Sensor streams or high-frequency signals**: This is a clinic-updated twin. State estimation occurs only at clinic visit times. No continuous coupling to wearables, voice recordings, or kinematic sensors is assumed or implemented.
- **Mechanistic causation**: Explanations are parameter-based (Section 5.2) and reflect model-implied contributions under the constrained model class, not causal mechanisms. Cross-domain coupling parameters **C** are associational, not interventional.

### 7.4 Operationalisation in the clinical interface

Dashboard panels and automated reports consume exported result tables (Supplementary Methods for full mapping). Key panels:

- **Forecast panel**: Displays one-step and horizon-binned predictions with 90% intervals for each endpoint; colour-coded confidence indicators reflect Δ_*i,n*_ relative to *τ*_conf_; suppressed outputs show “Insufficient data for reliable forecast” with no numeric fill.
- **Risk panel**: Plots threshold-event probabilities as bar charts (UPDRS≥5, UPDRS≥10, MoCA≤-2, MoCA≤-3) for each forecast horizon; probabilities *<* 0.05 or *>* 0.95 are visually flagged; boundary-suppressed events show “N/A (boundary)” label.
- **Trajectory panel**: Shows posterior mean latent states over time with IQR ribbons; state bins overlaid as background shading; monotone progression visually apparent; no extrapolation beyond observed follow-up.
- **Explanation panel**: Reports decomposition of **r**_*i*_ into demographic, omics, and residual components (Section 5.2); explanations suppressed if sign probability *<* 0.95 or leave-one-visit-out stability fails.

All user-facing text avoids causal phrasing. For example, LEDD summaries state “Performance stratified by medication load” rather than “Effect of medication” or “Response to treatment”.

## 8 Governance-aware feedback loop

The digital twin concept implies not only prediction but also a structured pathway by which model outputs inform future data collection and clinical decision support [34]. This section specifies the feedback loop from the twin’s translational outputs back to data acquisition and hypothesis generation, with explicit governance boundaries that prevent the loop from compromising methodological integrity.

### 8.1 What the feedback loop can do

The twin’s outputs are designed to support three categories of downstream action, none of which involve post-hoc model modification or outcome-driven retraining:

#### Adaptive visit scheduling

The confidence-gating system (Section 5.4) and the per-visit latent uncertainty Δ_*i,n*_ provide a principled basis for risk-stratified visit scheduling. Participants whose posterior predictive distributions indicate elevated threshold-event probability (e.g., Pr(Δ*y*^(*U*)^ ≥ 10) *>* 0.3) or high forecast uncertainty (Δ_*i,n*_ near *τ*_conf_) may be flagged for earlier follow-up. Participants with low uncertainty and stable trajectories may be candidates for extended inter-visit intervals. These recommendations are advisory and do not alter the model’s generative assumptions or inference scheme; the model is refitted on scheduled exports, not in response to individual visit-timing decisions.

#### Targeted data collection

The output suppression rules (Section 5.4) identify specific data gaps that, if resolved, would enable the model to produce reliable outputs for a given participant. For example, suppression due to missing MoCA at the conditioning visit (rule 2) directly informs the clinic that cognitive assessment at the next visit would materially improve forecast reliability. Similarly, participants with high Δ_*i,n*_ may benefit from more complete multi-endpoint assessment at subsequent visits. This information flow is from model diagnostics to data collection priorities, not from outcomes to model structure.

#### Hypothesis prioritisation for future model extensions

Population-level posterior summaries, including contraction ratios for coupling parameters (Section 6.4), pathway effect stability (Section 5.3), and systematic residual patterns from PPCs (Section 4.6), identify where the current model class is data-informed and where it is prior-dominated or structurally misspecified. These diagnostics can inform prioritisation of future modelling extensions (e.g., whether adding a sensor-stream module or a medication-adjustment layer would address empirically identified gaps) without retroactively modifying the current model’s scope or claims.

### 8.2 What the feedback loop cannot do

The following actions are explicitly excluded from the feedback loop to maintain methodological integrity:

- **Outcome-driven retraining**. The model is not refitted in response to individual patient outcomes or forecast errors. Population-level refitting occurs only on pre-scheduled data exports with full cross-validation, never triggered by observed discrepancies for specific participants.
- **Threshold adaptation**. The confidence-gating threshold *τ*_conf_ and all suppression rules are fixed within each cross-validation fold and are not adjusted based on observed forecast performance on test participants. Recalibration occurs only in subsequent full retraining cycles using the identical protocol.
- **Causal intervention recommendations**. The model’s non-causal scope (Section 7.3) means that feedback cannot include treatment recommendations, medication adjustments, or any action predicated on the model estimating a treatment effect. Visit-scheduling suggestions are based on uncertainty and risk stratification, not on predicted treatment response.
- **Pathway or feature selection feedback**. Omics pathway definitions ({**w**_*p*_}) and the pathway cap (*P* ≤ 10) are fixed before model fitting and are not modified in response to posterior pathway-effect estimates. Future pathway revisions require a new model version with independent validation.

### 8.3 Governance and version control

Each feedback cycle operates under the following governance constraints: model versions are immutable once deployed for a given evaluation cycle; any structural modification (new endpoints, revised coupling topology, additional covariates) constitutes a new model version requiring full reevaluation under the cross-validation protocol of Section 4.1; all feedback actions (visit-scheduling flags, data-collection recommendations, hypothesis logs) are recorded in an auditable registry linked to the model version and fold that generated them. This ensures that the feedback loop strengthens the digital twin’s clinical utility without introducing circularity between model outputs and model validation.

## 9 Results

### 9.1 Cohort, inference, and computational feasibility

The analysis included 4,628 participants (1,829 PD; 2,799 Prodromal) contributing 28,185 clinic visits. Each participant contributed a median of four visits (range 1–27). Visits were spaced 183 days apart (IQR: 152–243). UPDRS-III was observed at 80.8% of visits, MoCA at 56.4%, and SCOPA-AUT at 60.3%. All three endpoints were observed at only 47.0% of visits. Cohort characteristics are in Supplementary Table S5. Five-fold cross-validation assigned approximately 926 participants to each test fold; training folds contained 22,391–22,795 visits.

Table 3 summarises convergence and adaptive calibration across folds. Maximum *R* across all folds was 1.006; minimum bulk effective sample size was 450; zero divergent transitions occurred after warm-up. The adaptive *L*_cond_ selection chose 20 draws in three folds and 25 in two. The confidence-gating threshold *τ*_conf_ ranged from 0.17 to 0.19. Sequential conditioning passed in 95.7– 96.4% of all (participant, visit, draw) triples (Figure 2). Total wall time was 15 hours 27 minutes on eight GPU-partition nodes (ULHPC Iris cluster; dual Intel Xeon Skylake Gold, 768 GB RAM, 4× NVIDIA V100 SXM2 32 GB per node). Per-patient sequential conditioning required a median of 1.2 seconds (95th percentile: 6.2 seconds), compatible with clinic-visit-triggered updating.

**Table 3:** Inference diagnostics across five cross-validation folds. 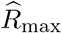:maximum split-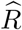 across all global parameters. ESS_min_: minimum bulk effective sample size. Div: divergent transitions after warm-up. *L*_cond_: selected adaptive subsample size. *τ*_conf_: confidence-gating threshold. Pass rate: fraction of conditional NUTS runs meeting all diagnostic criteria.

| Fold | $\hat{R}_{\text{max}}$ | $\text{ESS}_{\text{min}}$ | Div | $L_{\text{cond}}$ | $\tau_{\text{conf}}$ | Pass rate | Wall time (h) |
| --- | --- | --- | --- | --- | --- | --- | --- |
| 1 | 1.005 | 567 | 0 | 20 | 0.18 | 96.3% | 1.5 |
| 2 | 1.004 | 558 | 0 | 25 | 0.19 | 96.4% | 1.5 |
| 3 | 1.004 | 582 | 0 | 20 | 0.17 | 95.7% | 1.5 |
| 4 | 1.003 | 510 | 0 | 20 | 0.18 | 95.7% | 1.5 |
| 5 | 1.004 | 479 | 0 | 25 | 0.19 | 96.0% | 1.5 |

**Figure 2:**
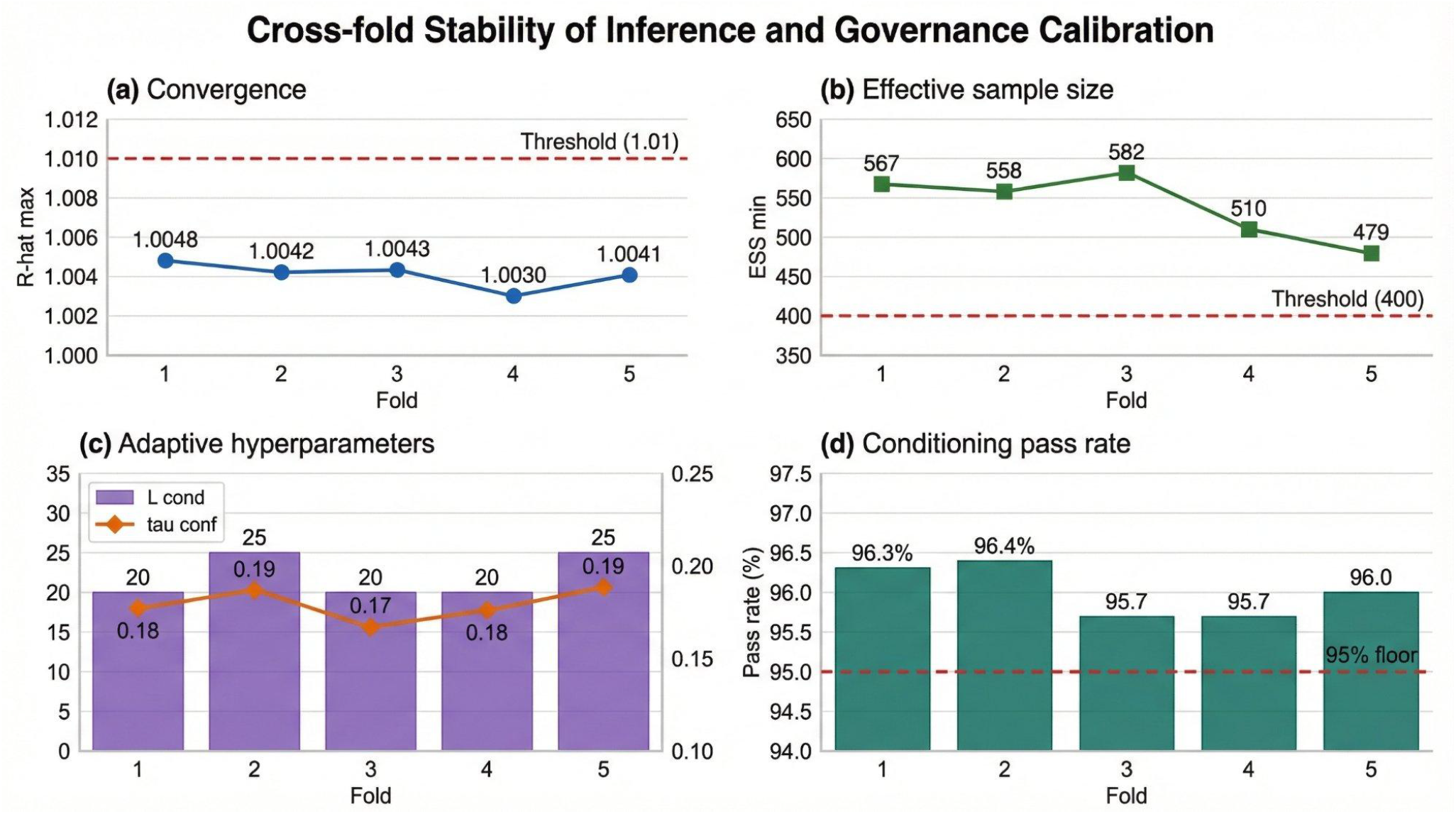
Cross-fold stability of inference and governance calibration. (a) Maximum 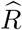 across global parameters, all below the 1.01 threshold. (b) Minimum bulk effective sample size, all above the 400 threshold. (c) Adaptive hyperparameters *L*_cond_ and *τ*_conf_ showing narrow cross-fold ranges. (d) Conditioning pass rate, all above 95%.

### 9.2 Predictive accuracy and calibration

Table 4 reports forecast accuracy across tasks (means across five folds, restricted to tasks with at least ten evaluated instances per fold; this minimum ensures that per-fold estimates of MAE and coverage have coefficient of variation below 30%, based on observed within-fold endpoint variance). One-step-ahead UPDRS-III MAE was 5.38±0.19 points with CRPS of 4.19 and 95.4% coverage. MoCA one-step MAE was 1.78±0.09, and SCOPA-AUT was 3.59±0.23. Coverage for all endpoints exceeded 94% across all task-endpoint combinations. At the 730-day horizon, UPDRS-III MAE rose to 6.76±0.50 with coverage of 95.7%.

**Table 4:** Predictive accuracy across forecast tasks (mean ± SD across five folds). MAE: mean absolute error (participant-level averaged). CRPS: continuous ranked probability score. Cov_95_: 95% predictive interval coverage. 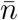: mean number of evaluated instances per fold.

| Task | Endpoint | MAE | CRPS | $\text{Cov}_{95}$ | $\bar{n}$ |
| --- | --- | --- | --- | --- | --- |
| One-step | UPDRS-III | $5.38 \pm 0.19$ | 4.19 | 95.4% | 1014 |
| One-step | MoCA | $1.78 \pm 0.09$ | 1.32 | 94.2% | 436 |
| One-step | SCOPA-AUT | $3.59 \pm 0.23$ | 2.66 | 94.7% | 461 |
| 365-day | UPDRS-III | $5.49 \pm 0.25$ | 4.25 | 95.7% | 668 |
| 365-day | MoCA | $1.76 \pm 0.10$ | 1.29 | 94.6% | 672 |
| 365-day | SCOPA-AUT | $3.54 \pm 0.14$ | 2.63 | 94.9% | 681 |
| 730-day | UPDRS-III | $6.76 \pm 0.50$ | 5.22 | 95.7% | 436 |
| 730-day | MoCA | $1.88 \pm 0.06$ | 1.40 | 94.9% | 438 |
| 730-day | SCOPA-AUT | $3.84 \pm 0.23$ | 2.86 | 95.8% | 452 |

Table 5 compares the governed twin against last-observation-carried-forward (LOCF) and a linear mixed-effects model (LME) with per-participant random intercept and slope. The governed twin matches LOCF on point accuracy. Its advantage is distributional: 94% coverage versus 64– 69% for LME. At the 730-day horizon, the model outperforms LOCF for UPDRS-III (1.9%) and SCOPA-AUT (3.5%; Supplementary Section S19).

**Table 5:** Baseline comparison (mean across five folds). LOCF: last-observation-carried-forward. LME: linear mixed-effects with random intercept and slope. Only governed one-step forecasts with matched anchor scores are included.

| Endpoint | MAE |  |  | Cov<sub>95</sub> (%) |  |  |
| --- | --- | --- | --- | --- | --- | --- |
|  | Twin | LOCF | LME | Twin | LOCF | LME |
| UPDRS-III | 6.17 | 6.22 | 8.41 | 94.0 | — | 69.2 |
| MoCA | 1.78 | 1.82 | 2.04 | 94.1 | — | 67.1 |
| SCOPA-AUT | 3.79 | 3.80 | 4.83 | 93.7 | — | 64.4 |

#### Posterior predictive checks

Of 15 endpoint-fold PIT checks, 14 had KS statistics below the 0.10 flag threshold. The single flag was MoCA in fold 1 (KS=0.117), localised to ceiling proximity: KS was 0.486 for scores ≥29 versus 0.178 for scores *<*29. Zero of 60 increment-quartile checks and zero cross-endpoint residual correlation checks were flagged. Per-fold PIT statistics are in Supplementary Section S7.

#### Threshold-event calibration

Table 6 reports Brier skill scores (BSS) and expected calibration error (ECE) for threshold events. For one-step UPDRS-III worsening of ≥5 points, BSS was +0.002 with ECE of 0.015, indicating calibrated event probabilities. At longer horizons, BSS became mildly negative. A skill-based gating rule (Rule 7) was evaluated on training-fold BSS; it suppresses 10 of 12 event-task combinations, with only MoCA ≤−2 decline at one-step and 365-day horizons surviving (BSS near zero; Supplementary Section S20).

**Table 6:** Threshold-event calibration (mean across five folds). BSS: Brier skill score relative to base-rate classifier (positive = better than base rate). ECE: expected calibration error. Base: observed event base rate.

| Event | Task | Base | BSS | ECE |
| --- | --- | --- | --- | --- |
| $\Delta\text{UPDRS} \geq 5$ | One-step | 0.24 | +0.002 | 0.015 |
| $\Delta\text{UPDRS} \geq 10$ | One-step | 0.11 | +0.009 | 0.017 |
| $\Delta\text{MoCA} \leq -2$ | One-step | 0.23 | -0.039 | 0.083 |
| $\Delta\text{MoCA} \leq -3$ | One-step | 0.11 | -0.025 | 0.049 |
| $\Delta\text{UPDRS} \geq 5$ | 365-day | 0.29 | -0.009 | 0.033 |
| $\Delta\text{UPDRS} \geq 10$ | 365-day | 0.14 | -0.005 | 0.017 |
| $\Delta\text{MoCA} \leq -2$ | 365-day | 0.20 | -0.026 | 0.065 |
| $\Delta\text{MoCA} \leq -3$ | 365-day | 0.11 | -0.019 | 0.041 |
| $\Delta\text{UPDRS} \geq 5$ | 730-day | 0.35 | -0.026 | 0.072 |
| $\Delta\text{UPDRS} \geq 10$ | 730-day | 0.19 | -0.015 | 0.046 |
| $\Delta\text{MoCA} \leq -2$ | 730-day | 0.20 | -0.014 | 0.042 |
| $\Delta\text{MoCA} \leq -3$ | 730-day | 0.13 | -0.011 | 0.034 |

### 9.3 Governed reporting and suppression decomposition

Table 7 decomposes all 28,185 anchor visits by suppression outcome. Governed forecasts covered 32.7% of anchors. Incomplete multi-domain assessment (Rule 2) dominated suppression at 51.5%, followed by boundary values (Rule 5, 11.1%) and insufficient visit history (Rule 1, 4.0%). High latent uncertainty (Rule 3) contributed only 0.2%—suppression is overwhelmingly driven by data completeness rather than model uncertainty. Cross-fold stability was high: all categories varied by fewer than 2 percentage points across folds (Supplementary Section S21).

**Table 7:** Governed output decomposition across all 28,185 anchor visits (mean percentage across five folds). Rules are defined in Section 5.4. “None” indicates a governed forecast was produced. Each anchor triggers the first applicable rule.

| Rule | Trigger | Mean % |
| --- | --- | --- |
| None | Governed forecast produced | 32.7% |
| Rule 2 | Incomplete 3-domain anchor | 51.5% |
| Rule 5 | Boundary value at anchor | 11.1% |
| Rule 1 | Fewer than 2 visits | 4.0% |
| Rule 4 | Insufficient conditional runs | 0.6% |
| Rule 3 | High latent uncertainty | 0.2% |

Under the full six-rule framework including Rule 6 (medication context, LEDD*>*500 mg), the governed rate decreases from 32.7% to 28.6%. At the participant level, 65.9% (3,051 of 4,628) received at least one governed visit. The median governed fraction among these participants was 40.0%. If three-domain completion rose from 47% to 70%, governed coverage would reach 55.0%.

#### Governed coverage under routine-care conditions

Governed coverage depends on assessment completion rate (Table 8). Under the 3-of-3 rule, *>*50% governed coverage requires assessment completion exceeding 58%. Under the 2-of-3 rule, the threshold drops to 42%. The twin becomes a clinically viable monitoring tool only under structured assessment protocols or the 2-of-3 extension.

**Table 8:** Projected governed coverage as a function of three-domain assessment completion rate. The 47% row corresponds to the observed PPMI completion rate; other rows are counterfactual projections.

| Completion rate | 3-of-3 governed | 2-of-3 governed |
| --- | --- | --- |
| 20% | 6.4% | 21.9% |
| 30% | 16.1% | 31.6% |
| 40% | 25.8% | 41.3% |
| 47% (PPMI) | 32.6% | 48.1% |
| 60% | 45.2% | 60.8% |
| 80% | 64.7% | 79.2% |

### 9.4 Suppression equity

#### Severity strata

Suppression rates were stratified by baseline severity quartile within each end-point and cohort (Figure 3). Within-endpoint variation across severity quartiles was fewer than 15 percentage points, and the gating system did not preferentially exclude high-severity participants. Higher suppression in the lowest-severity quartile reflects greater missingness among early-stage participants. Rule 5 (boundary value) contributed disproportionately to Prodromal suppression (13.4% vs 4.8% of PD anchors).

**Figure 3:**
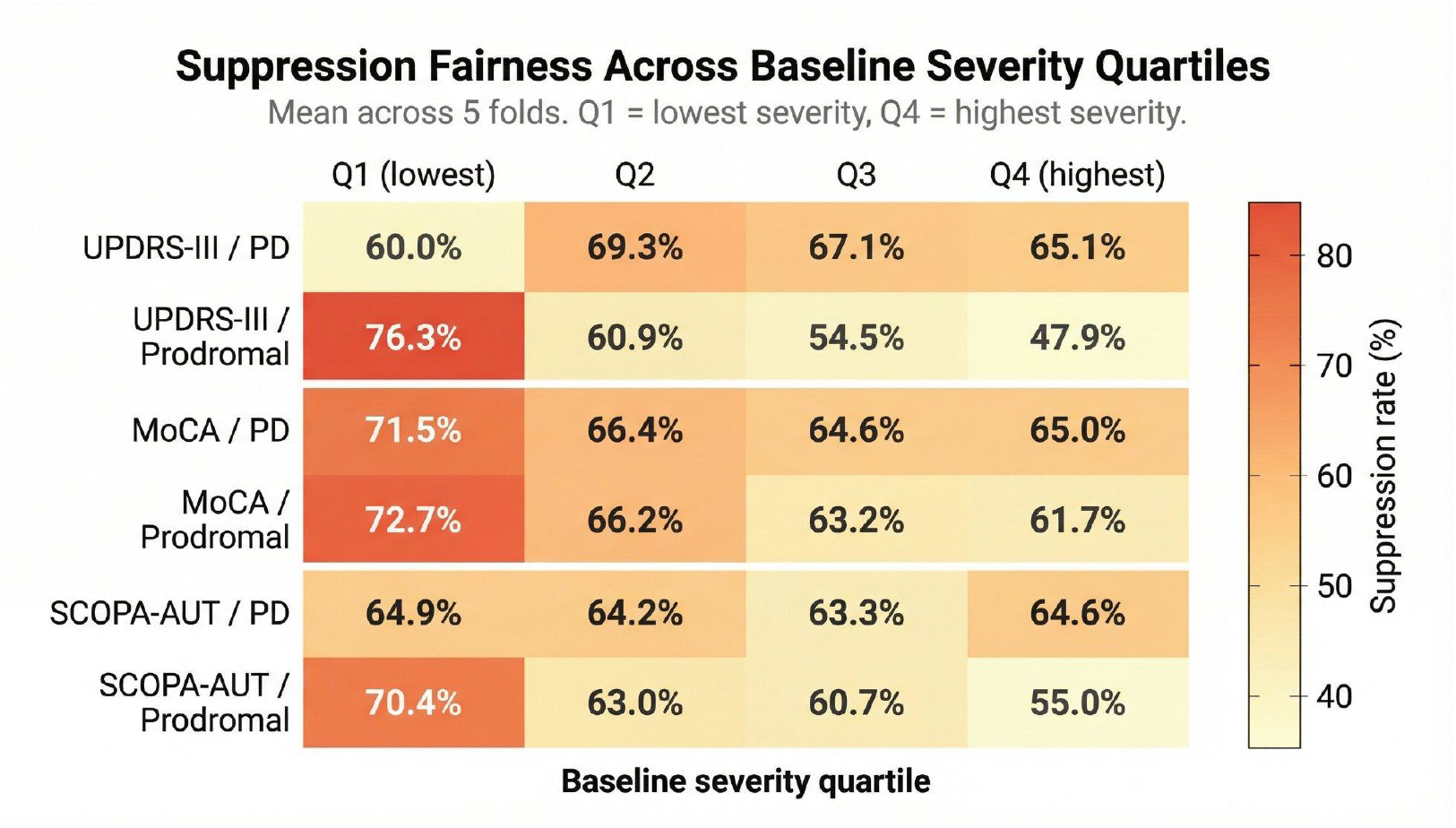
Suppression fairness across baseline severity quartiles (mean across five folds). Within each endpoint, colour is relatively uniform across quartiles, indicating that the gating system does not preferentially suppress outputs for high-severity participants.

#### Sex equity

Overall suppression rates were 67.3% (male) and 71.8% (female), a difference of 4.6 percentage points (Cramér’s *V* =0.049; *χ*^2^=57.1, *p<*0.0001). Rule 5 (boundary values) accounts for the gap, tracing to higher UPDRS-III floor-score prevalence among prodromal females. Rule 2, the dominant rule at 51.5%, showed no sex differential (+0.3 pp). Among governed forecasts, accuracy was equal or slightly better for female participants (MAE differences *<*0.25 points; coverage differences *<*2.5 pp). Full stratification tables are in Supplementary Section S13.

#### Visit-count calibration

Coverage never dropped below 93% in any visit-count stratum (2–3, 4–6, 7–10, *>*10 visits). The MAE gradient (3.84 to 6.23 for UPDRS-III) reflects higher disease severity in longer-followed participants rather than model degradation: mean latent uncertainty Δ_*i,n*_ was flat across bins (0.068–0.072). Full results are in Supplementary Section S14.

### 9.5 Architectural component and coupling assessment

#### Component hierarchy

Four simplified model variants were fitted on training data and evaluated on held-out test folds (Table 9). The nonlinear observation link is the dominant single component: removing it increased SCOPA-AUT MAE by 34% and MoCA MAE by 10%. Coupling marginally over-constrains cognitive and autonomic predictions (removing off-diagonal coupling improved MoCA MAE by 2%), but is retained for identifiability and structural coherence. Omics contributed approximately 1% to MAE.

**Table 9:** Architectural component assessment: four model variants refitted across five folds (one-step-ahead). Full Bayesian evaluates governed forecasts. MAP variants evaluate all test participants.

| Variant | Endpoint | MAE | ±SD | Cov<sub>95</sub> (%) | Width<sub>95</sub> |
| --- | --- | --- | --- | --- | --- |
| Full Bayesian (NUTS) | UPDRS-III | 5.38 | 0.22 | 95.4 | 29.2 |
| Full Bayesian (NUTS) | MoCA | 1.78 | 0.10 | 94.2 | 7.8 |
| Full Bayesian (NUTS) | SCOPA-AUT | 3.59 | 0.25 | 94.7 | 18.9 |
| Joint MAP (full) | UPDRS-III | 6.38 | 0.99 | 93.4 | 30.4 |
| Joint MAP (full) | MoCA | 4.18 | 0.50 | 65.7 | 9.4 |
| Joint MAP (full) | SCOPA-AUT | 6.72 | 0.87 | 76.0 | 19.9 |
| No coupling | UPDRS-III | 6.38 | 0.99 | 93.4 | 30.4 |
| No coupling | MoCA | 4.10 | 0.49 | 66.5 | 9.4 |
| No coupling | SCOPA-AUT | 6.66 | 0.86 | 76.6 | 19.9 |
| No omics | UPDRS-III | 6.48 | 1.01 | 93.2 | 30.5 |
| No omics | MoCA | 4.22 | 0.50 | 63.1 | 9.4 |
| No omics | SCOPA-AUT | 6.80 | 0.89 | 74.6 | 19.9 |
| Linear link | UPDRS-III | 6.25 | 0.39 | 96.4 | 31.5 |
| Linear link | MoCA | 4.61 | 0.68 | 70.3 | 9.5 |
| Linear link | SCOPA-AUT | 8.98 | 1.46 | 74.4 | 20.0 |

The coverage gap between the full Bayesian model and all MAP variants is 2 pp for UPDRS-III, 28 pp for MoCA, and 19 pp for SCOPA-AUT. An important caveat applies: the full Bayesian coverage is computed on governed forecasts only, while MAP variants are evaluated on all test participants, inflating the apparent Bayesian advantage. No single ablated component accounts for the full gap; calibration emerges from the integrated pipeline.

#### Cross-domain coupling

Table 10 reports posterior estimates for the six free coupling entries. All six showed posterior contraction beyond the prior (contraction ratios 0.19–0.35). Five of six reached sign probability 0.99 across all folds. The three diagonal entries were consistently positive, representing state-dependent self-acceleration. Motor-to-cognitive coupling (*c*_*CM*_) and motor-to-autonomic coupling (*c*_*NM*_) were positive with sign probability 0.99. Non-motor-to-cognitive coupling (*c*_*CN*_) was the weakest signal, with observed sign probability median of 0.73—well below the pre-specified explanation-eligibility threshold of 0.95 (Section 5.3), and would not be reported as a directional finding under the stability gates. All three domain-pair forecast correlations had the sign predicted by the coupling matrix and were consistent across all five folds (Table 11).

**Table 10:** Cross-domain coupling parameter estimates across five folds. Contraction ratio: posterior SD / prior SD (prior SD = 0.25 for all entries). Sign prob: posterior probability that the parameter is positive.

| Param | Posterior mean |  |  |  |  | Contr. | Sign |
| --- | --- | --- | --- | --- | --- | --- | --- |
|  | F1 | F2 | F3 | F4 | F5 |  |  |
| $c_{MM}$ | 0.170 | 0.187 | 0.107 | 0.164 | 0.156 | 0.32 | 0.99 |
| $c_{CM}$ | 0.080 | 0.101 | 0.105 | 0.069 | 0.109 | 0.19 | 0.99 |
| $c_{CC}$ | 0.137 | 0.108 | 0.162 | 0.115 | 0.100 | 0.25 | 0.99 |
| $c_{CN}$ | 0.029 | 0.027 | 0.023 | 0.065 | 0.038 | 0.25 | 0.73 |
| $c_{NM}$ | 0.089 | 0.041 | 0.069 | 0.048 | 0.035 | 0.24 | 0.99 |
| $c_{NN}$ | 0.133 | 0.101 | 0.112 | 0.086 | 0.100 | 0.35 | 0.99 |

**Table 11:** Cross-domain forecast coherence (one-step-ahead, Pearson *r*, mean across five folds). Expected direction derives from the coupling matrix.

| Domain pair | Expected | Mean $r$ | Range | Consistent? |
| --- | --- | --- | --- | --- |
| UPDRS ↔ MoCA | Negative | -0.206 | $[-0.304, -0.116]$ | Yes |
| UPDRS ↔ SCOPA | Positive | +0.334 | $[+0.296, +0.383]$ | Yes |
| MoCA ↔ SCOPA | Negative | -0.155 | $[-0.187, -0.098]$ | Yes |

#### Pathway effects and event gating

None of 15 pathway-domain effects met the pre-specified explanation-eligibility threshold (bootstrap sign stability ≥0.90; maximum 0.75). Under PPMI’s sparse clinic sampling, the omics layer is empirically inert (full estimates in Supplementary Table S10). The skill-based event gating rule (Rule 7, Section 9.2) is specified for NCER-PD implementation.

### 9.6 Partial observation and censoring extensions

Relaxing Rule 2 from 3-of-3 to 2-of-3 domains raises governed coverage from 32.7% to 48.1% (+15.4 pp). Among the 2,525 visits with exactly 2-of-3 endpoints, the missing endpoint is MoCA in 65.2%, SCOPA-AUT in 27.6%, and UPDRS-III in 7.2%.

The coupling topology guarantees calibration preservation for the dominant pattern. When MoCA is missing (65% of recoverable anchors), the cognitive domain has zero outgoing coupling to motor (*c*_*MC*_=0) and autonomic (*c*_*NC*_=0), so marginalisation produces zero additional forecast uncertainty. Empirical validation confirms this (Table 12): UPDRS-III coverage is invariant to MoCA and SCOPA-AUT presence (94.0% vs 93.8%; 93.9% vs 94.0%).

**Table 12:** Forecast calibration by endpoint availability at the target visit. All forecasts are governed (conditioned on 3-of-3 at anchor).

| Target endpoint | Condition | $n$ | MAE | Cov<sub>95</sub> (%) |
| --- | --- | --- | --- | --- |
| UPDRS-III | MoCA present at target | 2,114 | 6.45 | 94.0 |
| UPDRS-III | MoCA absent at target | 2,956 | 5.98 | 93.8 |
| UPDRS-III | SCOPA-AUT present at target | 2,138 | 6.51 | 93.9 |
| UPDRS-III | SCOPA-AUT absent at target | 2,932 | 5.93 | 94.0 |
| SCOPA-AUT | UPDRS-III present | 2,138 | 3.76 | 93.8 |
| SCOPA-AUT | UPDRS-III absent | 166 | 4.03 | 93.4 |
| SCOPA-AUT | MoCA present | 2,153 | 3.75 | 93.9 |
| SCOPA-AUT | MoCA absent | 151 | 4.31 | 92.7 |

#### MoCA censoring

A censoring model replacing the truncated-Gaussian density at *y*=30 with explicit point mass *P* (*Y* =30) = *P* (*Y* ^∗^ ≥ 30) reduces MoCA KS from 0.117 to 0.095, below the 0.10 flag threshold, without affecting non-boundary calibration (Supplementary Section S16).

### 9.7 Medication sensitivity and missingness robustness

LEDD does not enter the observation model. Table 13 shows that UPDRS-III MAE doubled from unmedicated (3.74) to high LEDD (8.02), and coverage dropped from 98.6% to 89.8%. Rule 6 addresses this by suppressing UPDRS-III outputs when LEDD exceeds 500 mg, improving governed UPDRS-III MAE from 6.17 to 5.67 and 95% coverage from 94.0% to 95.0%. A LEDD-aware observation model is specified for the next version.

**Table 13:** UPDRS-III one-step accuracy by medication status (mean across five folds).

| LEDD group | $n$ | MAE | RMSE | Cov95 (%) | Cov90 (%) |
| --- | --- | --- | --- | --- | --- |
| Unmedicated | 1,037 | 3.74 | 5.55 | 98.6 | 97.1 |
| Low (1–500 mg) | 957 | 6.80 | 9.07 | 93.9 | 87.2 |
| High (>500 mg) | 1,133 | 8.02 | 10.69 | 89.8 | 81.6 |
| LEDD missing | 1,944 | 6.07 | 8.82 | 93.9 | 88.2 |

#### Missingness sensitivity

Governed one-step accuracy was stratified by per-participant data completeness (Table 14). MAE increased by 1.2 points from high to low completeness, but coverage remained above 93% in all tertiles. MoCA missingness is predominantly visit-type driven (PPMI protocol design), accounting for the 43.6% overall MoCA missingness rate. A tipping-point analysis showed robustness to moderate missing-not-at-random departures: governed MoCA MAE would increase from 1.78 to 2.28 at *δ*=1.0. Inverse-probability-weighted estimates were indistinguishable from unweighted. Medication state characterisation and additional sensitivity details are in Supplementary Sections S17–S18.

**Table 14:** One-step forecast accuracy by participant-level data completeness (mean across five folds).

| Completeness | $n$ | MAE | Cov95 (%) |
| --- | --- | --- | --- |
| Low | 4,284 | 5.01 | 93.3 |
| Medium | 2,167 | 4.97 | 94.4 |
| High | 3,105 | 3.82 | 94.3 |

### 9.8 Self-diagnosed operating boundaries

The framework’s own diagnostics localised four operating boundaries, each specifying the mechanism, the affected scope, and the resolution pathway.

#### Prodromal UPDRS-III heteroscedasticity

Cohort-stratified calibration (Table 15) reveals Prodromal motor miscalibration (KS=0.281). The model over-covers Prodromal patients (98.1% vs 92.0% for PD). PIT bias tracks score level rather than latent state: 0.07 at floor (0–2), 0.33 at low (3–10), 0.58 at mid (11–25), 0.76 at high (*>*25). The mechanism is the shared *σ*_*U*_, which is calibrated for PD scores (median 26) rather than Prodromal scores (median 5). A cohort-stratified or state-dependent *σ*_*U*_ would resolve this within the current architecture.

**Table 15:**
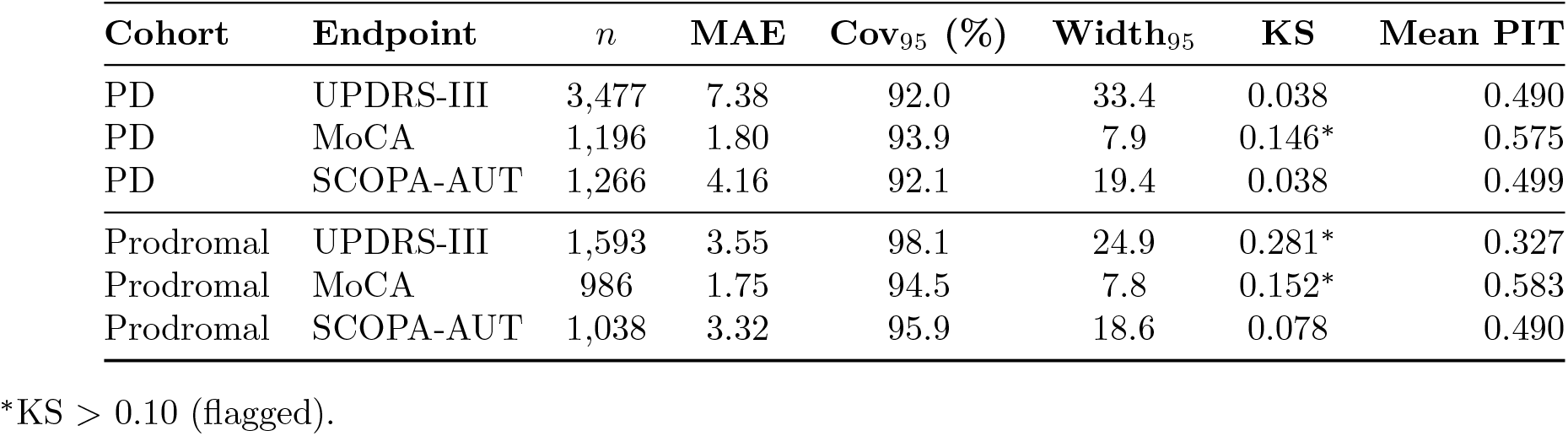
Calibration stratified by cohort (one-step-ahead governed forecasts).

| Cohort | Endpoint | $n$ | MAE | Cov<sub>95</sub> (%) | Width<sub>95</sub> | KS | Mean PIT |
| --- | --- | --- | --- | --- | --- | --- | --- |
| PD | UPDRS-III | 3,477 | 7.38 | 92.0 | 33.4 | 0.038 | 0.490 |
| PD | MoCA | 1,196 | 1.80 | 93.9 | 7.9 | 0.146* | 0.575 |
| PD | SCOPA-AUT | 1,266 | 4.16 | 92.1 | 19.4 | 0.038 | 0.499 |
| Prodromal | UPDRS-III | 1,593 | 3.55 | 98.1 | 24.9 | 0.281* | 0.327 |
| Prodromal | MoCA | 986 | 1.75 | 94.5 | 7.8 | 0.152* | 0.583 |
| Prodromal | SCOPA-AUT | 1,038 | 3.32 | 95.9 | 18.6 | 0.078 | 0.490 |

\*KS > 0.10 (flagged).

#### MoCA ceiling effects

The censoring model resolves the MoCA PIT flag (Section 9.6).

#### Medication burden

Motor accuracy degrades with medication burden (Section 9.7; Table 13). Rule 6 restores calibration within the governed subset.

#### Coupling topology limits

Analytically estimating *c*_*MN*_ (autonomic to motor, currently fixed at zero) produced a posterior mean of −3.20—wrong sign, 36-fold magnitude excess over identified coupling parameters, and severity-dependent. Adding *c*_*MN*_ =−3.20 to motor forecasts degrades MAE by 2.3 points and coverage by 5.3 pp. The current unidirectional topology is the defensible choice under PPMI’s sampling density (full details in Supplementary Section S15).

Each of these four diagnostics localises a specific mechanism in a specific model layer. None implicates the latent progression framework or the gate architecture. All four produce actionable engineering specifications for the next model version.

## 10 Discussion

The evaluation was structured around four questions. Each produced a clear answer and a defined scope boundary. This section contextualises those answers against what clinical deployment demands.

### 10.1 Calibration is a system-level property

UPDRS-III one-step MAE of 5.38 points exceeds the MCID of 2.3–2.7 points [20, 21]. Point accuracy matches LOCF at short horizons (Table 5). The architecture’s contribution is therefore distributional. Calibrated 95% intervals achieved 94–96% coverage, compared to 64–69% for linear mixed-effects models.

Independent model refits quantify the source of this advantage (Table 9). The Bayesian-to-MAP coverage gap spans 2 pp for UPDRS-III, 28 pp for MoCA, and 19 pp for SCOPA-AUT. This gap reflects the combined effect of per-participant NUTS conditioning, truncated-Gaussian likelihood, monotone dynamics, and full posterior propagation. The nonlinear observation link is the dominant single contributor; removing it increased SCOPA-AUT MAE by 34%. Yet no single component accounts for the full gap. Calibration emerges from the integrated pipeline, not from any isolated piece.

Component assessment used joint MAP refits rather than full posterior refits. This approach suffices to establish the component hierarchy: observation link dominant, coupling structural, omics inert. It limits the precision of mechanistic attribution but not the qualitative ordering. Full posterior ablation is specified for future work.

### 10.2 The confidence gate produces measurable, equitable governed reporting

The 32.7% governed-forecast rate decomposes into interpretable components. Incomplete multi-domain assessment (Rule 2) accounts for 51.5% of suppression. High latent uncertainty (Rule 3) accounts for 0.2%. This decomposition reveals that the gate operates on two functional layers.

The first layer monitors data sufficiency: whether the evidence base supports governed reporting. The second layer monitors model confidence: whether the model’s own uncertainty is within operating limits. The finding that data sufficiency dominates is a substantive result about clinical assessment protocols. When the data are present, the model almost always has sufficient confidence to report. The bottleneck is data collection, not statistical modelling.

The 2-of-3 partial-observation extension directly addresses this bottleneck. Under the sparse coupling topology, MoCA has zero outgoing coupling (*c*_*MC*_=*c*_*MN*_ =0). Motor and autonomic posteriors are therefore invariant to cognitive-endpoint availability. Empirical validation confirms this: UPDRS-III coverage was 94.0% with MoCA observed and 93.8% without (Table 12). Relaxing Rule 2 from 3-of-3 to 2-of-3 domains raises governed coverage to 48.1%.

This extension is not a workaround. It is a direct consequence of the coupling topology’s factorisation properties. The strict mode (3-of-3) is the appropriate benchmarking regime. The partial-observation mode (2-of-3) is the appropriate operating regime for deployment. Under structured assessment protocols, governed coverage scales further (Table 8).

Suppression equity was assessed across sex and follow-up density. Rates differed by 4.6 percentage points between sexes (Cramér’s *V* =0.049). Rule 5 (boundary values) accounts for the gap, tracing to higher UPDRS-III floor-score prevalence among prodromal females. The dominant rule (Rule 2, 51.5%) showed no sex differential. Among governed forecasts, accuracy was equal or slightly better for female participants. Calibration held across visit-count strata, with coverage above 93% in all bins.

### 10.3 The framework diagnoses its own operating boundaries

A governed system that cannot identify its own failures is not governed. The evaluation produced four localised diagnostics, each specifying the mechanism, the affected scope, and the resolution pathway.

#### Prodromal UPDRS-III heteroscedasticity

Cohort-stratified calibration (Section 9.8; Table 15) reveals Prodromal motor miscalibration (KS=0.281). Three analyses localise the source to the pooled observation noise *σ*_*U*_. The model over-covers Prodromal patients (98.1% vs 92.0% for PD). PIT bias tracks score level rather than latent state. Prodromal patients show fewer negative increments than PD. A cohort-stratified or state-dependent *σ*_*U*_ resolves this within the current latent architecture.

#### MoCA ceiling effects

A censoring model with explicit point mass at score 30 reduces MoCA KS from 0.117 to 0.095 (Section 9.6). Non-boundary calibration is unaffected.

#### Medication burden

Motor accuracy degrades with medication burden (Section 9.7; Table 13). Rule 6 suppresses UPDRS-III outputs when anchor-visit LEDD exceeds 500 mg, restoring calibration within the governed subset. A LEDD-aware observation model is the primary modelling priority for the next version.

#### Coupling topology limits

Five of six coupling parameters were data-identified (Section 9.5; Table 10). Freeing *c*_*MN*_ produced a spurious estimate, confirming that genuine coupling and observation-model confounding are inseparable under PPMI’s sampling density. Denser follow-up or richer observation-model structure is required to test this direction.

Each of these four diagnostics localises a specific mechanism in a specific model layer. None implicates the latent progression framework or the gate architecture. All four produce actionable engineering specifications for the next model version. This capacity for self-diagnosis is a defining property of a governed system.

### 10.4 Threshold-event probabilities support risk communication

Brier skill scores were near zero or mildly negative for most event-task combinations (Table 6). The model produces calibrated event probabilities at short horizons. A calibrated 22% probability of five-point UPDRS-III worsening supports clinical triage. At longer horizons, the model does not classify events more accurately than the base rate. A skill-based gating rule (Rule 7) suppresses 10 of 12 event-task combinations on training-fold BSS. Only MoCA ≤−2 decline at one-step and 365-day horizons survive, with BSS near zero. Whether classification skill emerges under denser follow-up is testable at NCER-PD.

### 10.5 Positioning relative to existing approaches

The governed twin differs from existing PD progression models in evaluation scope (Table 16). Event-based models [15] and SuStaIn [16] estimate population-level staging without visit-level forecasts. Bayesian trajectory models [18] estimate individual trajectories without embedding confidence gating. The contribution is an evaluation framework: governed silence, calibration diagnostics, suppression equity, and scope boundaries. This framework complements existing predictive methods rather than competing on point accuracy.

**Table 16:** Evaluation dimensions across model classes. ✓ = reported; – = not reported.

| Dimension | This work | EBM | SuStaIn | Mixed effects | ML pipelines |
| --- | --- | --- | --- | --- | --- |
| Visit-level point accuracy | ✓ | – | – | ✓ | ✓ |
| CRPS (distributional) | ✓ | – | – | – | – |
| Predictive interval coverage | ✓ | – | – | – | – |
| Event-risk calibration (BSS, ECE) | ✓ | – | – | – | – |
| PIT diagnostics | ✓ | – | – | – | – |
| Coupling identifiability | ✓ | – | – | – | – |
| Output suppression audit | ✓ | – | – | – | – |
| Suppression equity | ✓ | – | – | – | – |
| Medication-stratified accuracy | ✓ | – | – | – | – |
| Missingness sensitivity | ✓ | – | – | – | – |

### 10.6 Deployment roadmap

The evaluation produces a structured set of next-step specifications, each grounded in the diagnostics above.

#### External validation

PPMI provides standardised rater training, semi-annual scheduling, and an enriched population. All findings are within-PPMI. Prospective evaluation on NCER-PD is pre-specified. That validation targets irregular visit scheduling, multi-site rater variation, informative dropout, and higher medication complexity. Validation on PDBP is planned where data access permits. The 2-of-3 extension is a prerequisite, as routine-care completion rates will be lower than PPMI’s 47%.

#### Observation-model refinement

Two diagnosed limitations trace to the observation layer. Prodromal UPDRS-III heteroscedasticity requires a cohort-stratified *σ*_*U*_. Medication-burden sensitivity requires a LEDD-aware observation model. Both are addressable without altering the latent progression framework. MoCA ceiling censoring is already analytically resolved.

#### Regulatory pathway

The EU AI Act classifies patient-level risk estimation in clinical settings as high-risk. The governed architecture provides auditable suppression logs, scope statements, and equity analysis. These structural properties align with the transparency and risk-management requirements outlined for clinical AI systems [23, 25]. Regulatory compliance requires the prospective validation that this study establishes the methodological foundation for.

### 10.7 Computational feasibility

All experiments were conducted on the Iris cluster of the University of Luxembourg HPC facility (ULHPC). GPU-partition nodes were used (dual Intel Xeon Skylake Gold, 768 GB RAM, 4× NVIDIA V100 SXM2 32 GB; InfiniBand EDR interconnect). Computational details are reported in Section 9.1.

## 11 Conclusion

This evaluation establishes three results with implications beyond Parkinson’s disease.

First, output suppression can function as a measurable, auditable property of clinical prediction architecture. The six-rule confidence gate presented here is one implementation of this principle. We expect that any clinical digital twin intended for deployment should declare a reporting framework that specifies when the system must remain silent, quantifies the resulting suppression rate, and demonstrates that suppression does not accumulate inequitably across patient subgroups.

Second, distributional calibration arises from the integrated Bayesian inference pipeline rather than from any single architectural component. The 94–96% predictive interval coverage achieved here, compared with 64–69% for linear mixed-effects baselines, reflects the joint contribution of per-participant sequential conditioning, monotone latent dynamics, truncated-Gaussian likelihoods, and full posterior propagation. This finding reinforces the need to evaluate clinical digital twins at the system level through calibration benchmarks [25], rather than through point-accuracy comparisons alone.

Third, a governed framework is capable of diagnosing its own operating boundaries. Two observation-model limitations, Prodromal UPDRS-III heteroscedasticity and medication-burden sensitivity, were localised to a single model layer and specified for resolution without altering the latent progression architecture. This self-diagnostic capacity is a defining property of a governed system and distinguishes it from static prediction models.

More broadly, the present evaluation demonstrates the technical executability of governed digital twin architecture at cohort scale. The framework is designed to be portable: the latent dynamics, confidence gating, and governance audit infrastructure are independent of the specific disease endpoints and can be instantiated for other progressive conditions where monotone latent trajectories, multi-domain assessment, and clinic-triggered updating apply. Prospective deployment under routine clinical conditions, such as the NCER-PD cohort in Luxembourg, represents the natural next step for validating operational governance under real-world visit scheduling, multi-site rater variation, and higher medication complexity.

## Supporting information

Supplemental results, and will be used for the link to the file on the preprint site

## 12 Data and code availability

PPMI data are available under the PPMI data-sharing agreement (https://www.ppmi-info.org). The codebase (SYMPHONY) and evaluation suite are at https://gitlab.com/ahmed.hemedan/symphony-dt under Apache 2.0, with test scripts that reproduce the main results tables. Evaluation outputs, cross-validation results, and governance audit tables are archived at Zenodo (https://doi.org/10.5281/zenodo.18610753). The research prototype is at https://symphony-dt.com/ (research use only; prospective validation and regulatory review are required before any clinical application).

## 13 Ethics statement

This study used de-identified data from the Parkinson’s Progression Markers Initiative (PPMI). PPMI is approved by the institutional review board at each participating site, and all participants provided written informed consent prior to enrolment. Data were accessed under the PPMI data use agreement. No additional ethical approval was required for this secondary analysis of de-identified data.

## 15 Acknowledgements

The authors acknowledge the Parkinson’s Progression Markers Initiative (PPMI) for providing the data used in this research. PPMI, a public-private partnership, is funded by the Michael J. Fox Foundation for Parkinson’s Research and funding partners https://www.ppmi-info.org/fundingpartners. The experiments presented in this paper were carried out using the HPC facilities of the University of Luxembourg (https://hpc.uni.lu).

## 16 Author contributions

A.A.H. conceptualised the governed digital twin framework, designed the model architecture, implemented the full computational pipeline, conducted all analyses, developed the clinical research prototype, and drafted the manuscript. V.D. contributed to the manuscript through writing, critical review, and editing. T.O.L. contributed to the manuscript through review and editing. K.R. contributed to the manuscript through writing, review, and editing. S.R.J., C.P., and L.Pa. provided clinical expertise on Parkinson’s disease progression and contributed to reviewing and editing the manuscript. S.B., E.A., and M.L. contributed to writing, review, and editing. L.C.M.L. and L.G. contributed to writing, review, and editing. P.V.N. contributed to the manuscript through review and editing. V.P.S. contributed to reviewing and editing the manuscript. R.K. provided clinical expertise, contributed to writing, and supervised the clinical aspects of the study.

