## Supplemental results, and will be used for the link to the file on the preprint site for "A clinic-updated digital twin for Parkinson’s disease progression: Governed Bayesian forecasting with uncertainty-gated reporting"

**2 S2. Cohort characteristics** **6**

**3 S3. Full prior specification with rationale** **7**

**4 S4. Coupling topology and mechanistic rationale** **8**

**5 S5. Per-fold global parameter estimates** **9**

**6 S6. Pathway definitions and identifiability** **11**

**10 S10. Evaluation eligibility counts** **16**

**11 S11. Leakage control specification** **17**

**12 S12. 2-of-3 domain partial observation projection** **17**

**13 S13. Sex-stratified suppression equity** **18**

**14 S14. Calibration by visit count** **19**

**15 S15. Coupling topology sensitivity** **19**

**16 S16. MoCA censoring model** **20**

**17 S17. Cohort-stratified calibration and monotone-constraint assessment** **21**

**18 S18. Missingness sensitivity and computational requirements** **22**

**19 S19. LOCF comparison across horizons** **23**

**20 S20. Skill-based event gating (Rule 7)** **24**

**21 S21. Per-fold governed output decomposition** **25**

### 1 S1. Data preprocessing and harmonisation

#### 1.1 S1.1 Data source

Longitudinal data were obtained from the Parkinson's Progression Markers Initiative (PPMI), a multicentre prospective biomarker study [1, 2]. PPMI enrols participants across four cohorts: Parkinson's Disease, Prodromal, Healthy Control, and SWEDD. All analyses use the data export with cut date 28 January 2026. PPMI clinic visits are nominally semi-annual over an initial five-year window, subsequently extended. The present extract spans a maximum of 15.2 years from baseline.

#### 1.2 S1.2 Cohort selection

Participants were selected using the `COHORT_DEFINITION` field. Parkinson's Disease and Prodromal cohorts were retained. Healthy Controls ($n$=400) and SWEDD ($n$=81) were excluded because the monotone latent constraint is structurally incompatible with non-degenerative trajectories. The APPRDX code was available for 834 of 4,628 included participants (18.0%); no discrepancies with `COHORT_DEFINITION` were observed.

#### 1.3 S1.3 Individual eligibility

Participants required non-missing sex, non-missing birth date, and at least one clinic visit in the master visit index. No participants meeting the cohort criteria were excluded for missing covariates.

#### 1.4 S1.4 Visit structure and time alignment

A master visit index was constructed as the union of all (participant, event, date) tuples across the three outcome tables. Per participant-visit pair, the canonical visit time is the earliest calendar month among assessment records sharing the same event identifier, anchored to day 15. For 426 of 28,185 visits (1.5%), outcome forms within the same event spanned two calendar months; the earliest month determines the canonical time. Per-participant baseline is the earliest date in this index. Time since baseline is measured in days.

#### 1.5 S1.5 Clinical outcome derivation

**MDS-UPDRS Part III** ($y^{(U)}$, support $[0, 132]$). Extracted from NP3TOT. The raw export contains 36,336 rows for 4,830 participants; 34.4% are paired on-/off-medication records. Duplicates were resolved by preferring the row with non-missing NP3TOT, breaking ties by earliest assessment time. The strategy selects by completeness rather than medication state to ensure consistency with routine documentation.

**MoCA** ($y^{(Q)}$, support $[0, 30]$). Extracted from MCATOT. No duplicate records were identified. MoCA missingness (43.6%) substantially exceeds UPDRS-III missingness (19.2%) because not all PPMI visit types include cognitive assessment.

**SCOPA-AUT** ($y^{(S)}$, support $[0, 69]$). Derived by summing 25 item columns (SCAU1–SCAU25), each on $\{0, 1, 2, 3\}$. A valid total required at least 20 of 25 non-missing items. No item imputation was performed.

#### 1.6 S1.6 LEDD harmonisation

LEDD was harmonised from the Concomitant Medication Log following the official PPMI methodology [3], using conversion factors consistent with Tomlinson et al. [4]. Per-visit LEDD sums active medication records at the visit month. Missing stop dates are treated as ongoing. LEDD does not enter the likelihood or latent dynamics. Among 39.7% of visits with active medication ($n$=11,190), median LEDD was 500 mg/day (IQR: 300–800).

#### 1.7 S1.7 Omics preprocessing

Genetic consensus data are available for 3,345 of 4,628 participants (72.3%). Only pre-baseline draws are eligible. Participants with no pre-baseline draw are assigned $\mathbf{z}_i=\mathbf{0}$. Pathway scoring, standardisation, and feature selection are performed within training folds at model-fitting time to prevent leakage. Pathway dimensionality is capped at $P \leq 10$.

#### 1.8 S1.8 Outcome distributions and boundary prevalence

Table S1 reports distributional summaries. MoCA ceiling prevalence (14.2% at score 30) and UPDRS-III floor prevalence (10.3% at score 0) are the primary boundary considerations.

Table S1: Observed outcome distributions in the analysis dataset. Statistics computed on observed (non-missing) values only.

| **Outcome** | $n$ | **Mean** | **SD** | **Median** | **Range** | **Floor %** | **Ceiling %** |
|---|---|---|---|---|---|---|---|
| UPDRS-III | 22,765 | 17.0 | 14.8 | 15 | [0, 100] | 10.3% | 0.0% |
| MoCA | 15,883 | 26.7 | 3.0 | 27 | [0, 30] | 0.0% | 14.2% |
| SCOPA-AUT | 16,996 | 11.7 | 7.4 | 10 | [0, 55] | 1.2% | 0.0% |

#### 1.9 S1.9 Date-anchoring conventions

Table S2 specifies the deterministic day-anchoring rule for each field type. All PPMI dates recorded at month resolution are mapped to a fixed day before temporal computation.

Table S2: Date-anchoring conventions applied uniformly to all temporal fields.

| **Field type** | **Day anchor** | **Rationale** |
|---|---|---|
| Visit dates | Day 15 | Mid-month anchor minimises expected error in interval computation |
| Birth dates | Day 15 | Consistent with visit anchoring for age computation |
| Medication start | Day 1 | Conservative: avoids false non-coverage |
| Medication stop | Day 28 | Conservative: missing stop dates treated as ongoing |

#### 1.10 S1.10 Stepwise attrition

Table S3 reports the filtering sequence from the raw PPMI export to the final analysis dataset.

#### 1.11 S1.11 Outcome availability by visit

Table S4 reports visit-level outcome availability across the full dataset.

Table S3: Stepwise attrition from raw export to analysis dataset.

| **Stage** | **Excluded** | **Remaining** | **Reason** |
|---|---|---|---|
| Raw PPMI export | — | 5,109 | Starting pool |
| Exclude Healthy Controls | 400 | 4,709 | Non-degenerative trajectories |
| Exclude SWEDD | 81 | 4,628 | Non-degenerative trajectories |
| Missing sex or birth date | 0 | 4,628 | Baseline covariates required |
| Age outside [18, 110] years | 0 | 4,628 | Implausible age filter |
| No clinic visits in master index | 0 | 4,628 | At least one visit required |
| **Analysis dataset** | — | **4,628** | 1,829 PD; 2,799 Prodromal |

Table S4: Outcome availability across 28,185 clinic visit rows.

| **Pattern** | **Visits** | **%** |
|---|---|---|
| UPDRS-III observed | 22,765 | 80.8% |
| MoCA observed | 15,883 | 56.4% |
| SCOPA-AUT observed | 16,996 | 60.3% |
| All three observed | 13,245 | 47.0% |
| At least one observed | 24,645 | 87.4% |
| All three missing | 3,540 | 12.6% |

#### 1.12 S1.12 Pipeline output files

The preprocessing pipeline writes five deterministic output files: baseline table (4,628 rows), visit table (28,185 rows), all-cohorts baseline table (5,109 rows), all-cohorts visit table (31,677 rows), and omics table (3,345 rows). All pipeline decisions are recorded in a machine-readable audit file.

#### 2 S2. Cohort characteristics

Table S5: Cohort summary. LEDD statistics restricted to visits with at least one active medication (39.7% of visit rows). IQR = interquartile range.

| **Characteristic** | **Value** | **Note** |
|---|---|---|
| *Participants* | | |
| Total, $N$ | 4,628 | 1,829 PD; 2,799 Prodromal |
| Sex (male), $n$ (%) | 2,491 (53.8%) | |
| Sex (female), $n$ (%) | 2,137 (46.2%) | |
| Age at baseline, mean (SD) | 65.6 (8.4) years | |
| Age at baseline, median [IQR] | 66.4 [61.4, 71.1] | |
| Age at baseline, range | 20.8–92.6 years | |
| *Clinic visits* | | |
| Total visit rows | 28,185 | Unique (participant, event) pairs |
| Visits per participant, median (range) | 4 (1–27) | |
| $N_i \geq 2$ visits, $n$ (%) | 3,509 (75.8%) | |
| $N_i \geq 3$ visits, $n$ (%) | 3,029 (65.4%) | |
| >5 years follow-up, $n$ (%) | 923 (19.9%) | Max: 15.2 years |
| Inter-visit interval, median [IQR] | 183 [152, 243] days | |
| *Optional omics* | | |
| Pre-baseline omics draw, $n$ (%) | 3,345 (72.3%) | |

### 3 S3. Full prior specification with rationale

Table S6 lists every prior used in the model with its distributional form, scale, and scientific rationale. Priors are weakly informative by design. Sensitivity to prior scales is assessed via contraction ratios (Section S5).

Table S6: Prior specification. All priors are fixed before model fitting and do not adapt to data.

| **Parameter** | **Prior** | **Rationale** |
|---|---|---|
| $\boldsymbol{\mu}_r$ | $\mathcal{N}(\mathbf{0}, \mathbf{I}_3)$ | Centres population mean progression drive near zero; wide enough to accommodate PD and Prodromal cohorts |
| $\mathrm{vec}(\boldsymbol{\Gamma})$ | $\mathcal{N}(\mathbf{0}, 0.5^2\mathbf{I})$ | Moderate shrinkage on demographic effects; age and sex shifts are expected to be small relative to within-patient variation |
| $A_{j,p}$ | $\mathcal{N}(0, 0.25^2)$ | Strong shrinkage on pathway effects reflecting small expected marginal genetic contributions [5] |
| $c_{jk}$ (free) | $\mathcal{N}(0, 0.25^2)$ | Equivalent shrinkage on coupling; limits cross-domain influence to be data-driven |
| $\sigma_{r,\cdot}$ | HalfNormal(0.5) | Allows moderate between-participant heterogeneity in progression rates |
| $\sigma_{\xi,\cdot}$ | HalfNormal(0.15) | Centres process variability near zero; regularises toward smooth progression where visit-to-visit latent change is small |
| $a_k$ | $\mathcal{N}(0, 1)$ | Offset parameter for observation link; centred at midpoint of logit scale |
| $b_k$ | LogNormal$(0, 0.5^2)$ | Slope of observation link; positivity enforced; median 1.0 allows flexible mapping |
| $\sigma_k$ | HalfNormal$(0.1(U_k - L_k))$ | Observation noise scaled to 10% of endpoint range; $\sigma_U$=13.2, $\sigma_Q$=3.0, $\sigma_S$=6.9 |
| $\mathbf{s}_i(t_{i,0})$ | $\mathcal{N}(\mathbf{0}, \mathbf{I}_3)$ | Initial latent state centred at logit-midpoint; allows full range of baseline severity |

### 4 S4. Coupling topology and mechanistic rationale

The coupling matrix $\mathbf{C}$ has fixed sparse topology with six free entries and three fixed zeros:

$$\mathbf{C} = \begin{pmatrix} c_{MM} & 0 & 0 \\ c_{CM} & c_{CC} & c_{CN} \\ c_{NM} & 0 & c_{NN} \end{pmatrix}.$$

The topology prevents cyclic feedback and limits degrees of freedom. Zero entries ($c_{MC}$, $c_{MN}$, $c_{NC}$) enforce the prior assumption that motor impairment may influence cognitive and autonomic progression but not vice versa, consistent with Braak staging [6]. The diagonal entries represent state-dependent self-acceleration. The off-diagonal entries encode cross-domain influence: $c_{CM}$ (motor severity accelerates cognitive decline [7]), $c_{NM}$ (motor severity accelerates autonomic decline [8]), and $c_{CN}$ (autonomic burden accelerates cognitive decline).

### 5 S5. Per-fold global parameter estimates

Table S7 reports posterior summaries for all global parameters across five folds. Contraction ratio $\rho = \mathrm{sd}_{\mathrm{post}}/\mathrm{sd}_{\mathrm{prior}}$; values below 0.50 indicate meaningful posterior contraction.

Table S7: Global parameter posterior summaries. Mean: posterior mean (fold 1). SD: posterior standard deviation (fold 1). CR: contraction ratio (fold 1). SP: posterior sign probability >0 (fold 1). Values for folds 2–5 are available in the code repository.

| **Parameter** | **Prior SD** | **Mean** | **SD** | **CR** | **SP** | $\widehat{R}$ | **ESS** |
|---|---|---|---|---|---|---|---|
| *Population mean drives* | | | | | | | |
| $\mu_{r,M}$ | 1.00 | −0.409 | 0.150 | 0.15 | 0.01 | 1.000 | 1050 |
| $\mu_{r,C}$ | 1.00 | −0.799 | 0.134 | 0.13 | 0.01 | 1.002 | 911 |
| $\mu_{r,N}$ | 1.00 | −0.333 | 0.069 | 0.07 | 0.01 | 1.001 | 1536 |
| *Demographic effects* | | | | | | | |
| $\gamma_{\mathrm{age},M}$ | 0.50 | 0.023 | 0.141 | 0.28 | 0.58 | 1.000 | 743 |
| $\gamma_{\mathrm{age},C}$ | 0.50 | 0.039 | 0.077 | 0.15 | 0.75 | 1.002 | 753 |
| $\gamma_{\mathrm{age},N}$ | 0.50 | 0.024 | 0.126 | 0.25 | 0.60 | 1.002 | 640 |
| $\gamma_{\mathrm{sex},M}$ | 0.50 | 0.084 | 0.126 | 0.25 | 0.83 | 1.000 | 2114 |
| $\gamma_{\mathrm{sex},C}$ | 0.50 | −0.032 | 0.108 | 0.22 | 0.35 | 1.001 | 1832 |
| $\gamma_{\mathrm{sex},N}$ | 0.50 | 0.016 | 0.108 | 0.22 | 0.58 | 1.001 | 1120 |
| *Coupling parameters* | | | | | | | |
| $c_{MM}$ | 0.25 | 0.170 | 0.079 | 0.32 | 0.99 | 1.001 | 1817 |
| $c_{CM}$ | 0.25 | 0.080 | 0.047 | 0.19 | 0.99 | 1.001 | 1727 |
| $c_{CC}$ | 0.25 | 0.137 | 0.064 | 0.25 | 0.99 | 1.001 | 1390 |
| $c_{CN}$ | 0.25 | 0.029 | 0.063 | 0.25 | 0.73 | 1.000 | 823 |
| $c_{NM}$ | 0.25 | 0.089 | 0.059 | 0.24 | 0.99 | 1.001 | 883 |
| $c_{NN}$ | 0.25 | 0.133 | 0.087 | 0.35 | 0.99 | 1.001 | 503 |
| *Variance components* | | | | | | | |
| $\sigma_{r,M}$ | 0.50 | 0.268 | 0.051 | 0.10 | 0.99 | 1.000 | 1708 |
| $\sigma_{r,C}$ | 0.50 | 0.233 | 0.079 | 0.16 | 0.99 | 1.000 | 2065 |
| $\sigma_{r,N}$ | 0.50 | 0.271 | 0.084 | 0.17 | 0.99 | 1.000 | 1449 |
| $\sigma_{\xi,M}$ | 0.15 | 0.059 | 0.007 | 0.05 | 0.99 | 1.000 | 1708 |
| $\sigma_{\xi,C}$ | 0.15 | 0.050 | 0.019 | 0.12 | 0.99 | 1.000 | 1114 |
| $\sigma_{\xi,N}$ | 0.15 | 0.054 | 0.014 | 0.09 | 0.99 | 1.001 | 1925 |
| *Observation model* | | | | | | | |
| $a_U$ | 1.00 | −1.197 | 0.149 | 0.15 | 0.01 | 1.001 | 698 |
| $a_Q$ | 1.00 | −0.608 | 0.150 | 0.15 | 0.01 | 1.002 | 1397 |
| $a_S$ | 1.00 | −0.880 | 0.190 | 0.19 | 0.01 | 1.002 | 1776 |
| $b_U$ | 0.50 | 2.799 | 0.055 | 0.11 | 0.99 | 1.001 | 1403 |
| $b_Q$ | 0.50 | 2.106 | 0.160 | 0.32 | 0.99 | 1.001 | 1355 |
| $b_S$ | 0.50 | 2.412 | 0.065 | 0.13 | 0.99 | 1.001 | 763 |
| $\sigma_U$ | 13.20 | 8.105 | 0.721 | 0.05 | 0.99 | 1.000 | 636 |
| $\sigma_Q$ | 3.00 | 2.297 | 0.848 | 0.28 | 0.99 | 1.003 | 685 |

*Table S7 continued*

| Parameter | Prior SD | Mean | SD | CR | SP | $\widehat{R}$ | ESS |
|---|---|---|---|---|---|---|---|
| $\sigma_S$ | 6.90 | 4.814 | 1.144 | 0.17 | 0.99 | 1.003 | 602 |

147 Table S8 summarises cross-fold stability for all global parameters. The narrow ranges confirm
148 that posterior estimates are stable across data splits.

Table S8: Cross-fold summary of global parameter posterior means (median and range across five folds) and contraction ratios (median and range). All values rounded to three decimal places.

| Parameter | Posterior mean | | | Contraction ratio | | |
|---|---|---|---|---|---|---|
| | Median | Min | Max | Median | Min | Max |
| $\mu_{r,M}$ | −0.414 | −0.419 | −0.402 | 0.150 | 0.056 | 0.184 |
| $\mu_{r,C}$ | −0.799 | −0.844 | −0.768 | 0.135 | 0.061 | 0.185 |
| $\mu_{r,N}$ | −0.337 | −0.365 | −0.319 | 0.069 | 0.056 | 0.197 |
| $c_{MM}$ | 0.164 | 0.107 | 0.170 | 0.314 | 0.296 | 0.346 |
| $c_{CM}$ | 0.101 | 0.069 | 0.104 | 0.231 | 0.189 | 0.280 |
| $c_{CC}$ | 0.115 | 0.100 | 0.137 | 0.233 | 0.184 | 0.278 |
| $c_{CN}$ | 0.029 | 0.023 | 0.045 | 0.254 | 0.210 | 0.310 |
| $c_{NM}$ | 0.048 | 0.035 | 0.089 | 0.252 | 0.188 | 0.322 |
| $c_{NN}$ | 0.101 | 0.086 | 0.133 | 0.231 | 0.181 | 0.350 |
| $\sigma_U$ | 8.102 | 8.084 | 8.116 | 0.055 | 0.055 | 0.130 |
| $\sigma_Q$ | 2.294 | 2.289 | 2.310 | 0.283 | 0.115 | 0.295 |
| $\sigma_S$ | 4.809 | 4.792 | 4.823 | 0.166 | 0.132 | 0.167 |

### 6 S6. Pathway definitions and identifiability

#### 6.1 S6.1 Pathway membership

Table S9 defines the five pathways used as omics-derived prior covariates. All pathway definitions are fixed before model fitting and are not modified based on posterior estimates.

Table S9: Pathway definitions. Each pathway $p$ is a binary membership vector over the $G$ genetic consensus features. Coverage is 72.3% of analysis participants.

| $p$ | Pathway | Member features | $\|\mathbf{w}_p\|_0$ |
|---|---|---|---|
| 1 | Kinase signalling | LRRK2, VPS35, WGS, WES, GWAS | 5 |
| 2 | Lysosomal glucocerebrosidase | GBA, SANGER, SVs, WGS, WES | 5 |
| 3 | Mitochondrial QC | PRKN, PINK1, PARK7, PATHVAR_COUNT, RNASEQ_VIS | 5 |
| 4 | Synuclein trafficking | SNCA, APOE_e4, RNASEQ_VIS, IU_Fingerprint, RNASEQ | 5 |
| 5 | Mixed PD core | PATHVAR_COUNT, RNASEQ_VIS, LRRK2, GBA, APOE_e4 | 5 |

#### 6.2 S6.2 Pathway effect estimates and explanation eligibility

Table S10 reports posterior summaries for all 15 pathway-domain effects from fold 1. No entry met the explanation-eligibility threshold (bootstrap sign stability $\geq 0.90$). This result was consistent across all five folds.

Table S10: Pathway effect posterior summaries (fold 1). CR: contraction ratio. SP: sign probability >0. BSS: bootstrap sign stability from 200 resamples. Eligible: whether the entry meets the explanation-eligibility threshold.

| Parameter | Mean | CR | SP | BSS | Eligible |
|---|---|---|---|---|---|
| $A_{M,\text{kinase}}$ | −0.014 | 0.35 | 0.42 | 0.61 | No |
| $A_{M,\text{lysosomal}}$ | +0.015 | 0.38 | 0.58 | 0.73 | No |
| $A_{M,\text{mito}}$ | −0.033 | 0.38 | 0.33 | 0.64 | No |
| $A_{M,\text{synuclein}}$ | +0.003 | 0.71 | 0.51 | 0.74 | No |
| $A_{M,\text{mixed}}$ | +0.000 | 0.49 | 0.50 | 0.55 | No |
| $A_{C,\text{kinase}}$ | −0.010 | 0.40 | 0.45 | 0.62 | No |
| $A_{C,\text{lysosomal}}$ | −0.045 | 0.68 | 0.37 | 0.59 | No |
| $A_{C,\text{mito}}$ | −0.040 | 0.57 | 0.36 | 0.66 | No |
| $A_{C,\text{synuclein}}$ | +0.046 | 0.59 | 0.66 | 0.69 | No |
| $A_{C,\text{mixed}}$ | −0.006 | 0.69 | 0.48 | 0.64 | No |
| $A_{N,\text{kinase}}$ | +0.018 | 0.36 | 0.60 | 0.73 | No |
| $A_{N,\text{lysosomal}}$ | +0.017 | 0.43 | 0.58 | 0.59 | No |
| $A_{N,\text{mito}}$ | +0.021 | 0.63 | 0.57 | 0.67 | No |
| $A_{N,\text{synuclein}}$ | −0.060 | 0.62 | 0.31 | 0.53 | No |
| $A_{N,\text{mixed}}$ | −0.039 | 0.65 | 0.38 | 0.75 | No |

### 7 S7. Posterior predictive check details

#### 7.1 S7.1 PIT calibration per fold

Table S11 reports Kolmogorov-Smirnov statistics for the probability integral transform across all endpoint-fold combinations. The flag threshold is KS > 0.10.

Table S11: PIT calibration: KS statistics per endpoint and fold. Flagged entries (KS > 0.10) are marked with †.

| Endpoint | Fold 1 | Fold 2 | Fold 3 | Fold 4 | Fold 5 |
|---|---|---|---|---|---|
| UPDRS-III | 0.067 | 0.068 | 0.052 | 0.071 | 0.059 |
| MoCA | 0.117† | 0.057 | 0.083 | 0.085 | 0.058 |
| SCOPA-AUT | 0.038 | 0.034 | 0.040 | 0.044 | 0.068 |

#### 7.2 S7.2 Increment dispersion ratios per fold

Table S12 reports dispersion ratios (mean absolute residual / mean predicted SD) by inter-visit interval quartile. All 60 checks (3 endpoints × 4 quartiles × 5 folds) remained below 1.0. No flags were raised.

Table S12: Increment dispersion ratios by inter-visit interval quartile (fold 1). Ratios below 1.0 indicate that the monotone constraint does not produce anomalous residual inflation. All folds showed the same pattern.

| Endpoint | Q1 | Q2 | Q3 | Q4 |
|---|---|---|---|---|
| UPDRS-III | 0.749 | 0.773 | 0.766 | 0.716 |
| MoCA | 0.723 | 0.702 | 0.737 | 0.732 |
| SCOPA-AUT | 0.696 | 0.683 | 0.767 | 0.682 |

#### 7.3 S7.3 Dispersion ratio ranges across all folds

Across all five folds, dispersion ratios ranged from 0.68 to 0.89 for UPDRS-III, from 0.70 to 0.85 for MoCA, and from 0.68 to 0.89 for SCOPA-AUT. No ratio exceeded 0.90. Zero of 60 interval-quartile checks were flagged. Cross-endpoint residual correlation checks also produced zero flags across all folds.

### 8 S8. Per-fold suppression breakdowns

#### 8.1 S8.1 Suppression by rule per fold

Table S13 reports the governed-output decomposition for each fold independently. The pattern is stable: Rule 2 dominates in every fold (51.2–51.8%), and Rule 3 contributes fewer than 0.4% in every fold. These decompositions reflect Rules 1–5 (structural gating). Rule 6 (medication context, LEDD > 500 mg) additionally suppresses UPDRS-III governed outputs, reducing the overall governed rate from 32.7% to 28.6% (Section 9.11 of the main text).

Table S13: Governed-output decomposition per fold. Each row shows the percentage of test-fold anchor visits assigned to each suppression category.

| **Category** | **F1** | **F2** | **F3** | **F4** | **F5** |
|---|---|---|---|---|---|
| Governed (none) | 33.7% | 31.9% | 32.3% | 32.9% | 32.5% |
| Rule 2 (incomplete anchor) | 51.2% | 51.8% | 51.6% | 51.4% | 51.4% |
| Rule 5 (boundary) | 10.6% | 11.6% | 11.4% | 10.7% | 11.4% |
| Rule 1 (<2 visits) | 3.6% | 4.1% | 3.8% | 4.1% | 4.2% |
| Rule 4 (run failures) | 0.8% | 0.5% | 0.6% | 0.7% | 0.4% |
| Rule 3 (low confidence) | 0.2% | 0.1% | 0.4% | 0.2% | 0.1% |
| $n$ anchors | 5,794 | 5,390 | 5,710 | 5,742 | 5,549 |

#### 8.2 S8.2 Suppression fairness by cohort and severity quartile

Table S14 reports mean suppression rates (averaged across five folds) stratified by endpoint, cohort, and baseline severity quartile. Q1 is the lowest-severity quartile; Q4 is the highest. Within each endpoint, variation across severity quartiles is fewer than 15 percentage points. The pattern is consistent: lower-severity strata have slightly higher suppression due to greater missingness among early-stage participants.

Table S14: Mean suppression rate (%) by endpoint, cohort, and baseline severity quartile (averaged across five folds). Q1: lowest severity. Q4: highest severity.

| **Endpoint** | **Cohort** | **Q1** | **Q2** | **Q3** | **Q4** |
|---|---|---|---|---|---|
| UPDRS-III | PD | 60.0 | 69.3 | 67.1 | 65.1 |
| | Prodromal | 76.3 | 60.9 | 54.5 | 47.9 |
| MoCA | PD | 71.5 | 66.4 | 64.6 | 65.0 |
| | Prodromal | 72.7 | 66.2 | 63.2 | 61.7 |
| SCOPA-AUT | PD | 64.9 | 64.2 | 63.3 | 64.6 |
| | Prodromal | 70.4 | 63.0 | 60.7 | 55.0 |

For Prodromal participants assessed by UPDRS-III, suppression drops from 76.3% in Q1 to 47.9% in Q4. This gradient reflects the fact that Prodromal participants with lower motor scores have less complete multi-domain assessment. The model does not preferentially suppress forecasts for sicker patients.

### 9 S9. Translational layer specifications

#### 9.1 S9.1 Output suppression triggers

Table S15 lists all conditions under which outputs are suppressed, the affected outputs, and the user-facing message displayed by the clinical prototype.

Table S15: Output suppression triggers and user-facing messages.

| Trigger | Technical definition | Affected outputs | User message |
|---|---|---|---|
| Insufficient visits | $N_i < 2$ | All forecasts, risks, explanations | “At least 2 visits required” |
| Missing anchor | Any of $y^{(U)}, y^{(Q)}, y^{(S)}$ missing at anchor | All forecasts, risks | “Assessment incomplete at anchor” |
| High uncertainty | $\Delta_{i,n} > \tau_{\text{conf}}$ | All forecasts, risks | “High uncertainty in state estimation” |
| Run failures | $< \max\{10, \lceil 0.4L_{\text{cond}} \rceil\}$ runs pass | All forecasts, risks | “Insufficient inference runs” |
| Boundary value | $y^{(k)}_{i,n} \in \{L_k, U_k\}$ | Event probabilities for endpoint $k$ only | “Score at boundary” |
| High LEDD | LEDD > 500 mg at anchor visit | UPDRS-III forecasts and risks | “Motor forecast suppressed (high medication burden)” |
| Sign instability | Sign prob < 0.95 or LOO instability | Explanations for that quantity | Suppressed silently |

#### 9.2 S9.2 User-facing text templates

All user-facing text avoids causal phrasing and adheres to the non-causal scope boundary.

**Forecast panel.** “Predicted UPDRS-III at next visit: [mean] (90% interval: [q05, q95]). This forecast reflects expected progression under current trajectory, not treatment response.”

**Risk panel.** “Probability of UPDRS-III worsening by ≥5 points within 6 months: [prob]%. This is a model-implied risk, not a guaranteed outcome.”

**Suppression (missing outcome).** “Outputs suppressed: one or more assessments missing at anchor. Complete motor, cognitive, and autonomic assessment at next visit.”

**Suppression (high uncertainty).** “Forecast suppressed due to high uncertainty in state estimation. Additional visits would improve personalisation.”

**Suppression (boundary).** “Event probability not computed (baseline score at scale boundary).”

**Suppression (high LEDD).** "Motor forecast suppressed: levodopa-equivalent daily dose exceeds 500 mg. Motor-score variability under high medication burden is not modelled. Cognitive and autonomic forecasts remain available if other rules are satisfied."

**LEDD stratification.** "Performance stratified by levodopa-equivalent daily dose. LEDD is descriptive only and does not enter predictions."

#### 9.3 S9.3 Dashboard panel mapping

Table S16 maps each dashboard panel to its source data, transformation, and visual element.

Table S16: Dashboard panel specifications.

| **Panel** | **Source data** | **Transformation** | **Visual element** |
|---|---|---|---|
| Forecast (one-step) | Predictive means and quantiles | None | Line plot with 90% ribbon |
| Forecast (horizon) | Horizon-binned predictions | Group by horizon | Bar chart with error bars |
| Risk | Event probabilities | None | Horizontal bar chart |
| Trajectory | Posterior latent states | IQR ribbons | Multi-line plot |
| Confidence indicator | 95% coverage statistic | Traffic-light rule | Icon (green/yellow/red) |

### 10 S10. Evaluation eligibility counts

Table S17 reports the number of forecast instances available for metric computation at each task-endpoint combination. Instances require both a non-suppressed forecast and an observed target value.

Table S17: Evaluation-eligible instances per fold (mean across five folds). "OK with observed" counts instances where status is non-suppressed and the target endpoint is observed.

| **Task** | **Endpoint** | **F1** | **F2** | **F3** | **F4–5 range** |
|---|---|---|---|---|---|
| One-step | UPDRS-III | 1,098 | 921 | 993 | 1,000–1,058 |
| One-step | MoCA | 504 | 375 | 428 | 431–444 |
| One-step | SCOPA-AUT | 523 | 400 | 442 | 464–475 |
| 365-day | UPDRS-III | 723 | 602 | 661 | 668–685 |
| 365-day | MoCA | 731 | 598 | 665 | 674–690 |
| 365-day | SCOPA-AUT | 739 | 620 | 669 | 677–699 |
| 730-day | UPDRS-III | 487 | 375 | 419 | 425–476 |
| 730-day | MoCA | 487 | 375 | 422 | 430–475 |
| 730-day | SCOPA-AUT | 506 | 400 | 424 | 434–498 |
| 180-day | UPDRS-III | 397 | 352 | 377 | 363–402 |
| 180-day | MoCA | 6 | 4 | 9 | 5–6 |
| 180-day | SCOPA-AUT | 9 | 4 | 11 | 5–8 |

The 180-day horizon for MoCA and SCOPA-AUT yields fewer than 10 instances per fold. This is because the PPMI assessment schedule rarely includes cognitive and autonomic assessment at the 6-month intermediate visit. These bins are excluded from the main results as unreliable.

### 11 S11. Leakage control specification

The following operations are computed on the training fold only within each cross-validation split:

1. Omics feature standardisation moments ($\widehat{\mu}_g$, $\widehat{\sigma}_g$) and pathway SD ($\widehat{\sigma}_p$).
2. Bootstrap stability checks for pathway effects $A_{j,p}$ using $B$=200 patient-level bootstrap resamples.
3. Confidence-gating threshold $\tau_{\text{conf}}$ selected on training-fold anchors.
4. Adaptive subsample size $L_{\text{cond}}$ selected using predictive stability criteria on 50 training participants.
5. LEDD quartile boundaries for stratified reporting.

Test-fold participants are never used for any calibration, threshold selection, or standardisation computation. All five-fold splits are participant-level and stratified by cohort designation (PD vs. Prodromal).

### 12 S12. 2-of-3 domain partial observation projection

Visit-level endpoint completion across 28,185 visits: 3-of-3 at 46.4% ($n$=13,245), exactly 2-of-3 at 16.0% ($n$=2,525), exactly 1-of-3 at 26.0%, and 0-of-3 at 11.6%. The 2-of-3 eligible visits ($n$=9,854; 62.4%) represent a 16.0 pp increase over 3-of-3.

Among the 2,525 visits with exactly two endpoints, the missing endpoint distribution is: MoCA missing in 1,647 visits (65.2%), SCOPA-AUT missing in 697 visits (27.6%), UPDRS-III missing in 181 visits (7.2%). MoCA missingness is protocol-driven: PPMI visit types V02, V05, V07, V09, and R01 omit cognitive assessment by design.

Table S18: Governed coverage projection under relaxed Rule 2.

| **Metric** | **3-of-3** | **2-of-3** | **Change** |
|---|---|---|---|
| Visit eligibility | 47.0% | 62.4% | +15.4 pp |
| R2-rescuable fraction | — | 29.9% of R2 | — |
| Governed anchor rate | 32.7% | ∼48.1% | +15.4 pp |
| Participants ≥1 governed | 65.9% | ∼83% | ∼+17 pp |

The coupling topology guarantees calibration preservation for the dominant pattern. When MoCA is missing (65% of recoverable anchors), the cognitive domain has zero outgoing coupling to motor ($c_{MC}$=0) and autonomic ($c_{NC}$=0) under the sparse topology. Marginalisation over the missing cognitive state therefore produces zero additional forecast uncertainty for UPDRS-III and SCOPA-AUT predictions. When SCOPA-AUT is missing (28%), the autonomic domain has weak coupling to cognitive ($c_{CN}$=0.036, partially identified) and zero coupling to motor ($c_{MN}$=0). When UPDRS-III is missing (7%), motor-to-cognitive ($c_{CM}$=0.093) and motor-to-autonomic ($c_{NM}$=0.057) contribute marginalisation uncertainty; the maximum interval widening is below 0.2% for all affected domain pairs.

Table S19: Suppression rates by sex (anchor-level, pooled across five folds).

| Sex | Anchors | Governed | Gov.% | Supp.% |
|---|---|---|---|---|
| Male | 13,293 | 4,353 | 32.7% | 67.3% |
| Female | 10,264 | 2,890 | 28.2% | 71.8% |

### 13 S13. Sex-stratified suppression equity

Rate difference (M−F): −4.6 pp [95% CI: −5.8, −3.4]. Cramér's $V$=0.049 (negligible effect size). $\chi^2$=57.1, $p$<0.0001. The statistical significance reflects the large sample, not the disparity magnitude.

Suppression decomposition by rule: Rule 2 (incomplete anchor) shows +0.3 pp sex difference (negligible). Rule 5 (boundary values) shows −5.0 pp (females higher). Rules 3 and 4 show <0.1 pp difference. The sex gap is driven entirely by boundary-value prevalence in prodromal females.

Table S20: Sex-stratified one-step forecast accuracy (governed forecasts only, mean across five folds).

| Sex | Endpoint | $n$ | MAE | $\text{Cov}_{95}$ (%) | $\text{Width}_{95}$ |
|---|---|---|---|---|---|
| M | UPDRS-III | 3,126 | 5.47 | 95.5 | 29.6 |
| F | UPDRS-III | 1,944 | 5.24 | 95.3 | 28.5 |
| M | MoCA | 1,328 | 1.85 | 93.2 | 8.0 |
| F | MoCA | 854 | 1.67 | 95.7 | 7.6 |
| M | SCOPA-AUT | 1,398 | 3.59 | 94.6 | 18.9 |
| F | SCOPA-AUT | 906 | 3.59 | 95.0 | 18.8 |

Governed forecast accuracy is equal or better for female participants across all endpoints. Coverage differences are below 2.5 pp. The gating system does not hide poor female performance behind higher suppression.

Table S21: Suppression by sex and cohort (anchor-level).

| Sex | Cohort | Anchors | Gov.% | Supp.% |
|---|---|---|---|---|
| M | Parkinson's Disease | 8,325 | 32.1% | 67.9% |
| F | Parkinson's Disease | 5,111 | 29.6% | 70.4% |
| M | Prodromal | 4,968 | 33.9% | 66.1% |
| F | Prodromal | 5,153 | 26.7% | 73.3% |

The sex difference is larger in the Prodromal cohort (M−F = −7.2 pp) than in PD (−2.4 pp), consistent with higher floor-score prevalence among prodromal females.

Cross-fold stability: mean difference −4.6 pp (SD 0.8 pp; range −5.5 to −3.2 pp across five folds).

Participant-level governed coverage: 83.3% of males and 77.4% of females had ≥1 governed visit. PD cohort: 82.8% (M) vs 83.7% (F), sex-equivalent. Prodromal: 83.8% (M) vs 74.0% (F).

Table S22: Participant characteristics by visit-count bin.

| **Bin** | $N$ | **Med. visits** | **Med. follow-up (yr)** | **Mean $x_M$ (last)** |
|---|---|---|---|---|
| 2–3 | 981 | 3 | 1.1 | 0.076 |
| 4–6 | 1,177 | 5 | 2.1 | 0.135 |
| 7–10 | 450 | 8 | 3.5 | 0.216 |
| >10 | 901 | 16 | 8.5 | 0.235 |

### 14 S14. Calibration by visit count

Visit count is a strong proxy for disease duration and severity. The >10 bin has 8× longer follow-up and 3× higher latent motor severity than the 2–3 bin.

Table S23: UPDRS-III one-step accuracy by visit-count bin (mean ± SD across five folds).

| **Bin** | **MAE** | **Cov$_{95}$ (%)** | **Width$_{95}$** | $\Delta_{\text{IQR}}$ |
|---|---|---|---|---|
| 2–3 | 3.84±0.55 | 98.1 | 26.8 | 0.069 |
| 4–6 | 4.99±0.49 | 97.0 | 28.5 | 0.069 |
| 7–10 | 5.95±0.87 | 94.4 | 31.7 | 0.071 |
| >10 | 6.23±0.12 | 93.1 | 29.8 | 0.069 |

MAE increases with visit count; this reflects severity, not miscalibration. Coverage remains above 93% in all strata. Latent uncertainty ($\Delta_{\text{IQR}}$) is flat (0.068–0.072), confirming the MAE gradient reflects disease-stage heterogeneity rather than differential estimation quality. MoCA MAE varies by 0.29 points across bins. SCOPA-AUT varies by 0.76 points.

Suppression rates by bin: 2–3 visits 53.5%, 4–6 visits 67.3%, 7–10 visits 74.9%, >10 visits 70.2%. The 2–3 bin has the highest governed rate (46.5%) because short-follow-up participants have proportionally fewer incomplete anchors (R2: 36.2% vs 59–66% in other bins).

### 15 S15. Coupling topology sensitivity

Table S24: Estimated $c_{MN}$ (autonomic → motor) across five folds. Prior: $\mathcal{N}(0, 0.25^2)$.

| **Fold** | **Post. mean** | **Post. SD** | **Contr. ratio** | **Sign(>0)** |
|---|---|---|---|---|
| 1 | −3.243 | 0.077 | 0.308 | 0.000 |
| 2 | −3.131 | 0.084 | 0.336 | 0.000 |
| 3 | −3.146 | 0.080 | 0.318 | 0.000 |
| 4 | −3.241 | 0.072 | 0.287 | 0.000 |
| 5 | −3.220 | 0.082 | 0.328 | 0.000 |

The estimate is strongly negative (mean −3.20; cross-fold SD 0.054). Its magnitude is 36× the identified $c_{NM}$ (0.057). All existing coupling parameters fall in $[0.04, 0.16]$; an estimate of −3.20 is two orders of magnitude out of family.

**Severity-quartile decomposition.** The motor-residual-autonomic correlation depends on motor severity. In Q1 (lowest; $x_M \in [0.01, 0.04]$; predominantly prodromal): $r = +0.054$, $p$=0.06 (positive, non-significant). In Q2 ($x_M \in [0.04, 0.14]$): $r = -0.272$, $p$<0.001. In Q3 ($x_M \in [0.15, 0.25]$):

$r{=}-0.119$, $p{<}0.001$. In Q4 ($x_M \in [0.26, 0.71]$): $r{=}-0.127$, $p{<}0.001$. The positive Q1 trend is directionally consistent with the biological hypothesis but below statistical significance. The strong negative signals in Q2–Q4 reflect severity-dependent measurement noise producing a spurious association.

Partial correlation (motor residual vs $x_N$ controlling for $x_M$): $r{=}-0.117$, $p{<}0.001$. The association partially traces to shared severity progression.

**Cohort stratification.** Prodromal: $c_{MN}{=}-2.74$, contraction ratio 0.49 (approaching prior-dominated). PD: $c_{MN}{=}-3.01$, contraction ratio 0.38. The coupling hypothesis is weakest in the Prodromal subgroup where the biology is strongest.

**Forecast impact.** Adding $c_{MN}{=}-3.20$ increases one-step UPDRS-III MAE by 2.3 points and reduces coverage by 5.3 pp (mean across five folds).

### 16 S16. MoCA censoring model

MoCA ceiling prevalence: 11.6% at score 30 (observed data across five test folds).

Table S25: PIT calibration by MoCA boundary proximity (mean across five folds).

| **Stratum** | $n$ | **Mean PIT** | **KS statistic** |
|---|---|---|---|
| Boundary (≥29) | 506 | 0.900 | 0.739 |
| Non-boundary (<29) | 1,676 | 0.482 | 0.085 |
| Overall | 2,182 | — | 0.153 |

The PIT miscalibration is confined to the boundary region. Non-boundary MoCA is well-calibrated (mean PIT 0.482; KS 0.085).

The censoring model replaces the truncated-normal density at $y{=}30$ with explicit point mass:

$$P(Y{=}30) = P(Y^* \geq 30) = 1 - \Phi\left(\frac{29.5-\mu}{\sigma}\right) \Big/ \left[\Phi\left(\frac{30-\mu}{\sigma}\right) - \Phi\left(\frac{0-\mu}{\sigma}\right)\right],$$

where $Y^*$ is the underlying continuous score and $\Phi$ is the standard normal CDF.

Table S26: MoCA censoring model effect on PIT calibration.

| **Metric** | **Current model** | **Censoring model** |
|---|---|---|
| Mean PIT at $y{=}30$ | 1.000 | 0.953 |
| Overall KS | 0.117 | 0.095 |
| Below 0.10 flag threshold? | No | Yes |

The censoring model reduces KS from 0.117 to 0.095, resolving the single MoCA PIT flag. Non-boundary calibration is unaffected (KS 0.085). Coverage at ceiling observations ($y{=}30$) is 96.2% under the current model and would narrow slightly under the censoring model. Coverage away from ceiling ($y{<}28$) is 92.0% and unchanged.

### 17 S17. Cohort-stratified calibration and monotone-constraint assessment

#### 17.1 PIT by cohort, per fold

Table S27: PIT calibration by cohort (one-step, pooled across endpoints, per fold).

| Fold | PD $n$ | PD KS | Prodromal $n$ | Prodromal KS |
|---|---|---|---|---|
| 1 | 1,378 | 0.036 | 747 | 0.123 |
| 2 | 944 | 0.045 | 752 | 0.091 |
| 3 | 1,136 | 0.050 | 727 | 0.089 |
| 4 | 1,286 | 0.047 | 691 | 0.112 |
| 5 | 1,195 | 0.048 | 700 | 0.117 |

PD calibration passes in all five folds (KS range 0.036–0.050). Prodromal calibration is borderline: KS exceeds 0.10 in three of five folds. The Prodromal miscalibration concentrates in UPDRS-III.

#### 17.2 Score-level PIT decomposition (Prodromal UPDRS-III)

Table S28: Prodromal UPDRS-III PIT by observed score level ($n$=1,593).

| Score stratum | $n$ | Mean PIT | KS | $\text{Cov}_{95}$ (%) |
|---|---|---|---|---|
| Floor (0–2) | 467 | 0.071 | 0.807 | 100.0 |
| Low (3–10) | 722 | 0.329 | 0.390 | 99.9 |
| Mid (11–25) | 319 | 0.584 | 0.193 | 96.9 |
| High (>25) | 85 | 0.758 | 0.401 | 77.6 |

PIT increases monotonically with observed score level. This is the signature of observation-model heteroscedasticity, not a latent constraint violation. The shared $\sigma_U$ produces intervals that are too wide for low scores (100% coverage at floor) and too narrow for high scores (77.6% at >25).

#### 17.3 Observed score increments by cohort

Table S29: Consecutive-visit UPDRS-III score increments by cohort. Negative = score improvement (observed, not latent).

| Cohort | Pairs | Neg (%) | Zero (%) | Pos (%) | Mean | Median |
|---|---|---|---|---|---|---|
| PD | 7,123 | 41.1 | 6.3 | 52.6 | +0.96 | +1 |
| Prodromal | 3,466 | 33.4 | 22.9 | 43.7 | +0.72 | 0 |

Prodromal participants show fewer negative increments than PD (33.4% vs 41.1%). The monotone constraint is less stressed in the Prodromal group. This rules out the monotone constraint as the source of Prodromal UPDRS-III PIT miscalibration.

##### 17.4 Medication state at UPDRS-III assessment

Among 12,939 UPDRS-III observed visits: 54.1% had LEDD=0 and 45.9% had LEDD>0. Within-participant LEDD CV=0.40 (mean SD=115 mg; mean range=328 mg). PPMI assesses UPDRS-III in the practically-defined OFF state for PD participants. Prodromal participants are typically unmedicated.

#### 18 S18. Missingness sensitivity and computational requirements

Governed coverage was projected across a range of assessment completion rates using the observed relationship between Rule 2 suppression and three-domain completion in PPMI. Non-R2 suppression (Rules 1, 3, 4, 5) was held constant at 15.9% of anchors. The 2-of-3 extension was modelled as a 16 pp increase in effective completion.

Under routine-care conditions with ∼30% three-domain completion, 3-of-3 governed coverage projects to 16.1% and 2-of-3 to 31.6%. Clinical viability (>50% governed) requires completion rates above 42% (2-of-3) or 58% (3-of-3).

**Computational requirements.** Population-level NUTS inference: 70 minutes per fold on eight GPU-partition nodes of the ULHPC Iris cluster (dual Intel Xeon Skylake Gold, 768 GB RAM, 4× NVIDIA V100 SXM2 32 GB; InfiniBand EDR). Total: 5.8h for five folds. Sequential conditioning: 15 minutes per fold (1.0s per test participant). Full five-fold pipeline wall time: 15.4 hours. Single-component NUTS ablation variants (no coupling, no omics, linear link): estimated 5×5.8h ≈ 29h additional NUTS time, assuming similar convergence rates.

### 19 S19. LOCF comparison across horizons

Table S30: LOCF comparison across horizons (MAE, mean across five folds).

| **Horizon** | **Endpoint** | **Twin** | **LOCF** | **Δ%** |
|---|---|---|---|---|
| 365-day | UPDRS-III | 6.22 | 6.22 | 0.0 |
| 365-day | MoCA | 1.76 | 1.76 | −0.0 |
| 365-day | SCOPA-AUT | 3.54 | 3.54 | 0.0 |
| 730-day | UPDRS-III | 7.71 | 7.86 | +1.9 |
| 730-day | MoCA | 1.94 | 1.93 | −0.1 |
| 730-day | SCOPA-AUT | 4.05 | 4.20 | +3.5 |

### 20 S20. Skill-based event gating (Rule 7)

A seventh gating rule was evaluated: suppress threshold-event probabilities at horizons where the model does not outperform the base rate on training-fold data.

Table S31: Rule 7 assessment: BSS by event type and horizon (mean across five training folds). Events with BSS<0 are suppressed.

| **Event** | **Horizon** | $n$ | **Base rate** | **BSS** | **Rule 7** |
|---|---|---|---|---|---|
| UPDRS $\geq$5 increase | One-step | 625 | 0.266 | $-0.057$ | Suppress |
| UPDRS $\geq$5 increase | 365-day | 411 | 0.287 | $-0.093$ | Suppress |
| UPDRS $\geq$5 increase | 730-day | 274 | 0.391 | $-0.094$ | Suppress |
| UPDRS $\geq$10 increase | One-step | 625 | 0.122 | $-0.011$ | Suppress |
| UPDRS $\geq$10 increase | 365-day | 411 | 0.130 | $-0.038$ | Suppress |
| UPDRS $\geq$10 increase | 730-day | 274 | 0.217 | $-0.054$ | Suppress |
| MoCA $\leq -2$ decline | One-step | 266 | 0.240 | $+0.001$ | Report |
| MoCA $\leq -2$ decline | 365-day | 414 | 0.216 | $+0.006$ | Report |
| MoCA $\leq -2$ decline | 730-day | 275 | 0.247 | $-0.004$ | Suppress |
| MoCA $\leq -3$ decline | One-step | 266 | 0.138 | $-0.012$ | Suppress |
| MoCA $\leq -3$ decline | 365-day | 414 | 0.126 | $-0.008$ | Suppress |
| MoCA $\leq -3$ decline | 730-day | 275 | 0.148 | $-0.011$ | Suppress |

All UPDRS-III worsening events have negative BSS across all horizons: the model produces calibrated distributional forecasts but has no event-classification skill for motor worsening. The two surviving MoCA events have BSS near zero, indicating marginal skill. Rule 7 is specified for NCER-PD implementation.

### 21 S21. Per-fold governed output decomposition

Table S32: Governed output decomposition per fold (% of test-fold anchor visits). Cross-fold stability is high: all categories vary by fewer than 2 percentage points across folds.

| Fold | Governed | R2 | R5 | R1 | R4 | R3 |
|---|---|---|---|---|---|---|
| 1 | 33.7 | 51.2 | 10.6 | 3.6 | 0.8 | 0.2 |
| 2 | 31.9 | 51.8 | 11.6 | 4.1 | 0.5 | 0.1 |
| 3 | 32.3 | 51.6 | 11.4 | 3.8 | 0.6 | 0.4 |
| 4 | 32.9 | 51.4 | 10.7 | 4.1 | 0.7 | 0.2 |
| 5 | 32.5 | 51.4 | 11.4 | 4.2 | 0.4 | 0.1 |
| **Mean** | **32.7** | **51.5** | **11.1** | **4.0** | **0.6** | **0.2** |
